# Biallelic *TET2* mutation sensitizes to 5’-azacitidine in acute myeloid leukemia

**DOI:** 10.1101/2021.07.14.21259597

**Authors:** Friedrich Stölzel, Sarah E. Fordham, Wei-Yu Lin, Helen Blair, Claire Elstob, Devi Nandana, Brigitte Mohr, Leo Ruhnke, Desiree Kunadt, Claudia Dill, Daniel Allsop, Rachel Piddock, Emmanouela-Niki Soura, Catherine Park, Mohd Fadly, Thahira Rahman, Abrar Alharbi, Manja Wobus, Heidi Altmann, Christoph Röllig, Lisa Wagenführ, Gail L. Jones, Tobias Menne, Graham H. Jackson, Helen J. Marr, Jude Fitzgibbon, Kenan Onel, Manja Meggendorfer, Olaf Heidenreich, Torsten Haferlach, Sara Villar, Beñat Ariceta, Rosa Ayala Diaz, Felipe Prosper, Pau Montesinos, Joaquin Martinez-Lopez, Martin Bornhäuser, James M. Allan

**Author notes:** Joint lead authors (Friedrich Stölzel, Sarah E. Fordham, Wei-Yu Lin). Corresponding authors: James M Allan, Newcastle Centre for Cancer, Newcastle University, Newcastle Upon Tyne, UK. Friedrich Stölzel, Medizinische Klinik und Poliklinik I, Universitätsklinikum Dresden, TU Dresden, Dresden, Germany.

## Abstract

Precision medicine can significantly improve outcomes for cancer patients, but implementation requires comprehensive characterization of tumor cells to identify therapeutically exploitable vulnerabilities. Here we describe somatic biallelic *TET2* mutation (focal deletion and nonsense mutation) in an elderly patient with acute myeloid leukemia (AML) that was chemoresistant to anthracycline and cytarabine, but acutely sensitive to 5’-azacitidine (5’-Aza) hypomethylating monotherapy, resulting in long-term morphological remission (overall survival (OS) 850 days). Given the role of TET2 as a regulator of genomic methylation, we hypothesized that mutant *TET2* allele dosage affects response to 5’-Aza. Using an isogenic cell model system and an orthotopic mouse xenograft, we demonstrate that biallelic *TET2* mutations confer sensitivity to 5’-Aza compared to cells with monoallelic mutation. We subsequently identified 29 additional patients from the Study Alliance Leukemia biobank with chromosome 4 abnormalities and identified two further patients with complex biallelic *TET2* mutations, including one with trisomy 4, homozygosity across the long arm and an inactivating point mutation. We also screened patients recruited to the PETHEMA FLUGAZA phase 3 clinical trial and identified three patients with biallelic *TET2* mutations, two of whom had responded very well to single agent 5’-Aza (OS 767 and 579 days) despite having adverse risk AML and poor performance status. Our data argue in favor of using hypomethylating agents for chemoresistant disease or as first line therapy in patients with biallelic *TET2*-mutated AML and demonstrate the importance of considering mutant allele dosage in the implementation of precision medicine for cancer patients.

**Key Points:**

- Mutant *TET2* allele dosage affects response to 5’-azacitidine in acute myeloid leukemia *in vitro* and in a xenograft model.
- Our data highlight the importance for screening of biallelic mutations to predict response to therapy in acute myeloid leukemia.

## Introduction

Acute myeloid leukemia (AML) is the exemplar of how interrogation of the somatic genome has facilitated understanding of disease pathogenesis and led to the development of novel therapies and stratified treatment approaches for some disease sub-groups^1,2^. Despite some success, therapeutic options for the majority of AML patients are limited and outcome remains very poor, with a 5-year overall survival (OS) of 15%^3^. Subsequent refinement of the AML genome has revealed a plethora of somatically mutated genes, including *TET2* which has been associated with poor outcome^1,4^.

Given the ineffectiveness of standard chemotherapy for many patients and the resulting poor outcome it is essential to identify genetic vulnerabilities that can be therapeutically exploited using existing treatments and the expanding catalogue of new agents. To this end, we present data that biallelic *TET2* mutation in AML confer sensitivity to hypomethylating chemotherapy. The TET2 enzyme catalyzes DNA demethylation by converting 5’-methylcytosine to 5’-hydroxymethylcytosine, and loss or attenuation of TET2 function leads to a somatically acquired global genomic hypermethylation and transcriptional and phenotypic re-programming underpinning the development of a leukemia phenotype^5^. As such, it is mechanistically plausible that hypomethylating chemotherapy could be particularly effective in TET2 null AML. These data serve as a paradigm for clinical diagnostics incorporating comprehensive genomic analyses to implement first line therapies with a higher likelihood of response in patients with AML.

## Methods

### Patients

AML patients with an abnormal chromosome 4 (UPN01-UPN30) were from the Study Alliance Leukemia (SAL) AML registry biobank in Dresden (Germany) which was approved by the Institutional Review Board of the Medical Faculty of Dresden (Ethikkomission der TU Dresden, Medizinische Fakultat Dresden; IRB #EK98032010).

*TET2* mutant patients over 65 years of age with newly diagnosed *TET2* mutant AML (UPN31-80) were from the Programa para el Estudio de la Terapeutica en Hemopatias Malignas (PETHEMA) phase 3 FLUGAZA clinical trial (NCT02319135)^6^, which was approved by the Institutional Review Board of Hospital Le Fe, Valencia, Spain (Eudract=2014-000319-15). Written informed consent was received from all study participants.

### AML cell lines and culture

AML cell lines were obtained from DSMZ (Braunschweig, Germany) and maintained in complete medium (CM) [RPMI1640 with 10% FBS and 50µg/ml penicillin/streptomycin] at 37°C in a humidified 5% CO_2_ incubator. The identity of cell lines was confirmed by short tandem repeat profiling (NewGene, Newcastle University, UK) and cultures were regularly tested for mycoplasma using a MycoAlert kit (Lonza, Slough, UK).

### CRISPR-Cas9 *TET2*-targeting

CRISPR-Cas9 *TET2*-targeted HEL cells were generated using a one vector system (pLV-U6-gRNA/EF1a-puro-2A-Cas9-2A-GFP in lentiviral particles (Sigma-Aldrich, Dorset, UK)) with the sgRNA sequence 5’GTTTGGTGCGGGAGCGAGC3’ targeting *TET2* exon 6 (*TET2* transcript ID ENST00000540549.5). Lentiviral particles were incubated with HEL cells (MOI 2) in CM supplemented with 8µg/ml hexadimethrine bromide and centrifuged at 800g for 30 min at 32°C. Transduced cells were selected by culture in CM with 2µg/ml puromycin then cloned by plating in soft agar [CM with 0.2% agarose]. *TET2* mutation was confirmed by Sanger sequencing (exon 6) as described in supplemental methods.

### *In vivo* mouse model

Eight week-old male and female *Rag2*^*−/−*^*Il2rg*^*−/−*^ (129×Balb/c) mice^7^ were used for *in vivo* investigations (UK Home Office Project License PPL60/4552). For IF injection (on day 0), mice were anaesthetized and 5×10^5^ cells (HEL *TET2* monoallelic and HEL *TET2* biallelic in a 1:1 ratio) in 20µl CM were injected through the knee into the marrow cavity of the right femur. Mice were monitored for signs of disease progression and were euthanized if any tumors reached a diameter of 15mm, or prior to this point if any signs of animal suffering were observed (including but not limited to sustained weight loss of 15% of normal weight and reduced mobility). 5’-Aza was dissolved in sterile water and administered via intraperitoneal (IP) injection once daily starting on day 28 for 5 days at 5mg/kg. Control mice received sterile water only via IP administration. Mice were euthanized on day 35 and tissues (BM from both femurs, PB, spleen and any tumors) were harvested during post-mortem investigation. Genomic DNA was extracted using either a DNA Mini kit or DNA Micro kit (Qiagen).

### *TET2* allele-specific qPCR assay

A custom TaqMan SNP Genotyping assay (Applied Biosystems, CA, USA) was designed using probes that differentiate between intact wild-type (WT) *TET2* sequence (HEL *TET2* monoallelic clones) and *TET2* sequence with a 4bp deletion generated by CRISPR-Cas9 targeting (HEL *TET2* biallelic clones). qPCR reactions were setup in triplicate and consisted of genomic DNA (50ng), primers (forward: 5’-GTGAAGAGAAGCTACTGTGTTTGGT-3’, reverse: 5’-ACAATCACTGCAGCCTCACA-3’), fluorescent allele-specific probes (WT *TET2*: 5’-CCAGCTCGCTCCCG-3’-VIC, 4bp deleted *TET2*: 5’-TGGCCAGCTCCCG-3’-FAM) and TaqMan SNP genotyping mastermix (Applied Biosystems) according to manufacturer’s recommended volumes. Controls were prepared using a 1:1 mix of DNA extracted from HEL *TET2* monoallelic and HEL *TET2* biallelic cells. Thermal cycling (50°C 2 min, 95°C 10 min, followed by 40 cycles of 95°C 15 sec, 60°C 1 min) was performed using a 7300 Real Time PCR System (Applied Biosystems). Detected fluorescence for the two probes was converted to Ct values using SDS v1.4 software (Applied Biosystems) and a sample was considered positive if Ct was 38 or higher for either allele. In samples where only one allele was amplified in all replicates (due to complete domination of one cell population in the sample), a Ct of 38 was assigned to the non-amplified allele such that the sample could be included in the analysis. For all samples, adjusted ΔCt values (difference between WT *TET2* allele Ct and 4bp deleted *TET2* allele Ct, adjusted by subtracting ΔCt calculated from the control DNA with 1:1 allelic ratio) were converted to inverse Log_2_ values, such that a value of 1 indicated a 1:1 ratio between the two alleles (and hence the two cell populations in the sample). Inverse Log_2_ [ΔCt] values were compared between 5’-Aza-treated and VC-treated mice for each tissue type using the Mann-Whitney test.

Detailed methods for BM morphological assessment, cytogenetic analyses, nucleic acid preparation, SNP array genotyping, whole exome sequencing, RNA sequencing and differential gene expression analysis, methylation array and differential methylation analysis, mutation analysis (TET2, NPM1 and FLT3), western immunoblotting, cell proliferation, drug sensitivity and cloning efficiency assays are provided in supplemental Methods.

### Data sharing statement

For all raw data, please contact.

## Results

### AML index case with biallelic *TET2* mutation

We describe a male who presented with AML characterized by a t(4;12) translocation (46,XY,t(4;12)(q2?;q13)[12]/46,XY[10]) (Figure 1A) which was resistant to remission induction chemotherapy with daunorubicin and cytarabine (Ara-C) (Figure 1B-C). Moreover, the patient became pancytopenic and developed acute septicemia requiring intensive care. Following recovery, the patient was treated with single agent 5’-azacitidine (5’-Aza) monthly as palliation (Figure 1B), which unexpectedly resulted in complete morphological remission (CR) (Figure 1C). The patient remained in CR for 24 months prior to emergence of relapsed AML which was unresponsive to chemotherapy and the patient died 28 months after first diagnosis. Autopsy revealed subtotal AML BM infiltration and multiple extramedullary AML sites (Figure S1).

**Figure 1.**
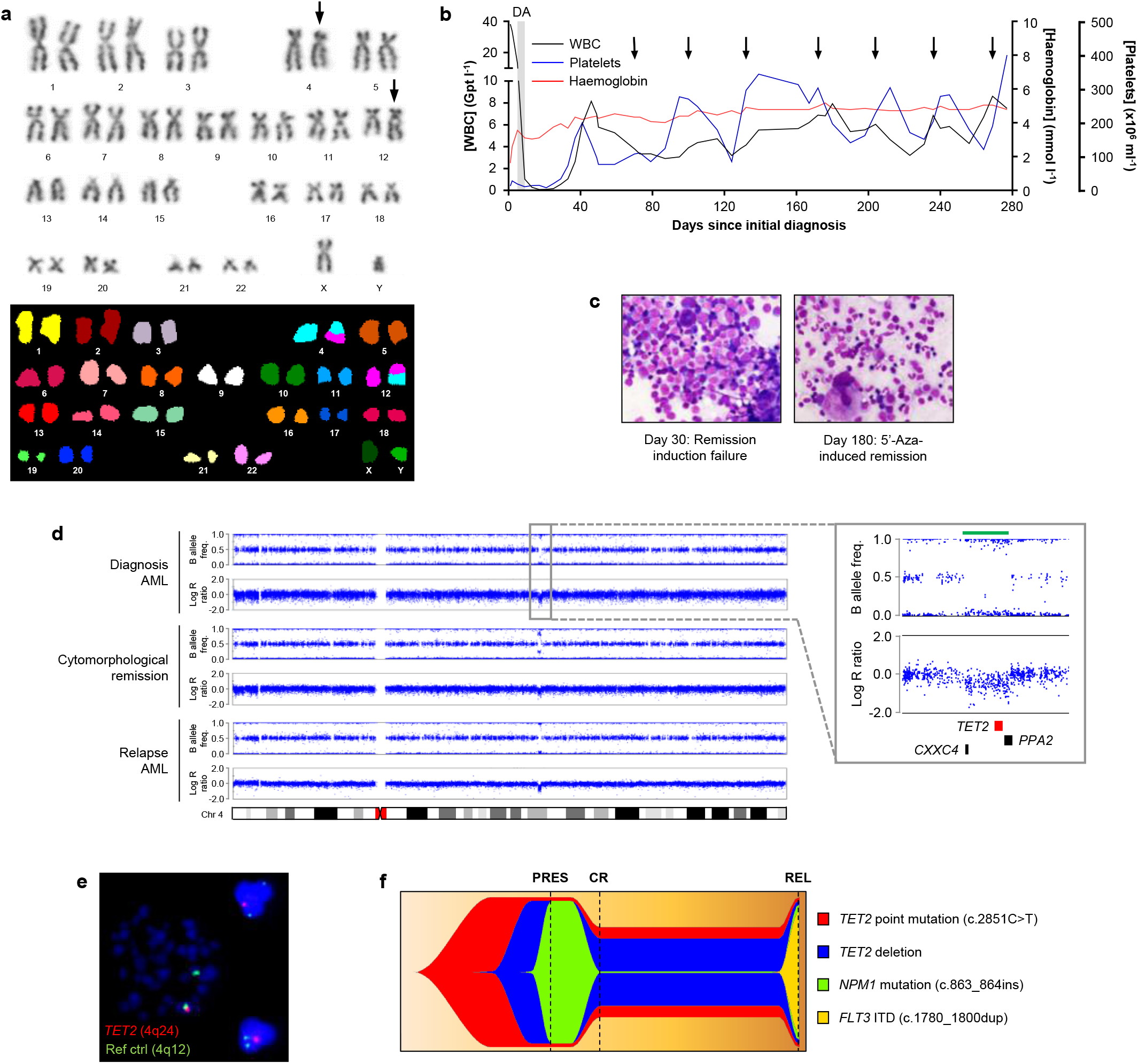
Biallelic *TET2* mutation in a patient with 5’-Aza-sensitive AML. (**a**) Identification of the t(4;12) translocation by G-banding (top) and spectral karyotyping (bottom) in leukemic blasts from the AML index patient (UPN01). Translocated chromosomes 4 and 12 are arrowed. (**b**) Hemoglobin, platelet and white blood cell (WBC) counts from diagnosis to relapse of UPN01. Grey shading indicates period of induction chemotherapy with daunorubicin and Ara-C (DA). Initiations of palliative 5’-Aza treatment cycles are arrowed. (**c**) Giemsa-stained BM smears at day 30 (left) (after failed induction chemotherapy) and day 180 (right) (during 5’-Aza-induced remission) in UPN01. Images are 500x magnification. (**d**) High density array copy number profiles of chromosome 4 from leukemic blasts of UPN01 at AML presentation, during CR and at relapse. Points represent individual SNPs, aligned relative to their position on chromosome 4 (indicated by the ideogram). Copy number is measured as Log R ratio, with 0 indicating diploid SNPs and positive and negative values indicating gain and loss, respectively. B allele frequency represents the ratio of the two alleles of each SNP such that 0.5 indicates allele heterozygosity and 0 and 1 indicate homozygosity. Inset shows expanded view of the boxed region. Green bar above plots highlights focal deletion within 4q24 encompassing *TET2, CXCC4* and *PPA2* (locations indicated by bars below plots). (**e**) FISH on leukemic blasts from UPN01 showing *TET2* (red) deletion in a metaphase cell (left of image) and two interphase cells (right of image). A probe binding within 4q12 (green) was used as reference. (**f**) Fishplot derived from sequencing analysis of leukemic blasts from UPN01 showing temporal acquisition of a *TET2* point mutation, *TET2* deletion, a *NPM1* insertion mutation and *FLT3* internal tandem duplication (ITD). Dashed lines represent timepoints at which blasts were analyzed. PRES; disease presentation, CR; complete remission, REL; relapse.

Single nucleotide polymorphism (SNP) array and FISH analysis of BM at AML presentation demonstrated that the major cell clone had a focal 1.1Mb deletion encompassing the *TET2, CXXC4* and *PPA2* genes (Figure 1D-E). Additionally, the retained *TET2* allele harbored a nonsense base substitution mutation in exon 3 (c.2815C>T, Q939*; Figure S2) which was detected in 96% of cells. The presentation AML was also characterized by a heterozygous *NPM1* mutation (c.863_864insTCTG; Figure S3). Construction of the tumor phylogeny indicated that disease pathogenesis was initiated by the *TET2* nonsense mutation with subsequent deletion of the second *TET2* allele, followed by acquisition of the *NPM1* mutation (Figure 1F). Both the *TET2* gene deletion and base substitution mutation were also present at high levels in the remission BM, despite this appearing morphologically normal (Figure 1C,F and Figure S2). The *NPM1* mutation was presumed to have persisted at below detection levels during remission given that it was a prominent feature of relapse disease (Figure 1F and Figure S2-3). The relapse was also characterized by a heterozygous *FLT3* internal tandem duplication (c.1780_1800dupTTCAGAGAATATGAATATGAT; Figure S4) which was not discernible in diagnostic or remission samples (Figure 1F and Figure S4).

These data demonstrate that 5’-Aza treatment almost completely eliminated the *TET2/NPM1*-mutated clone dominant at disease presentation. Although also reduced by 5’-Aza treatment, ancestral AML cells carrying biallelic *TET2* mutation, but negative for the *NPM1* mutation, retained viability and presumably re-acquired the ability to differentiate and recapitulate normal hematopoiesis rendering CM. Based on these observations, we hypothesized that mutant *TET2* allele dosage could affect cellular response and sensitivity to 5’-Aza.

### Biallelic *TET2* mutation result in a hypermethylation phenotype in AML cells and confer sensitivity to 5’-Aza hypomethylating chemotherapy *in vitro* and *in vivo*

In order to test whether biallelic *TET2* mutation sensitize AML cells to 5’-Aza we used CRISPR-Cas9 gene editing to completely inactivate *TET2* in the HEL AML cell line, which harbours a complex hypotriploid karyotype and a monoallelic *TET2* gene deletion (Figure 2A; Table S1)^8^. CRISPR-Cas9 was used to generate independent cell clones with deletions in *TET2* exon 6 (Figure S5) and null for TET2 protein expression (Figure 2B). Biallelic *TET2* mutation did not affect proliferation in liquid media (Figure 2C) nor cloning efficiency (CE) (Figure 2D).

**Figure 2.**
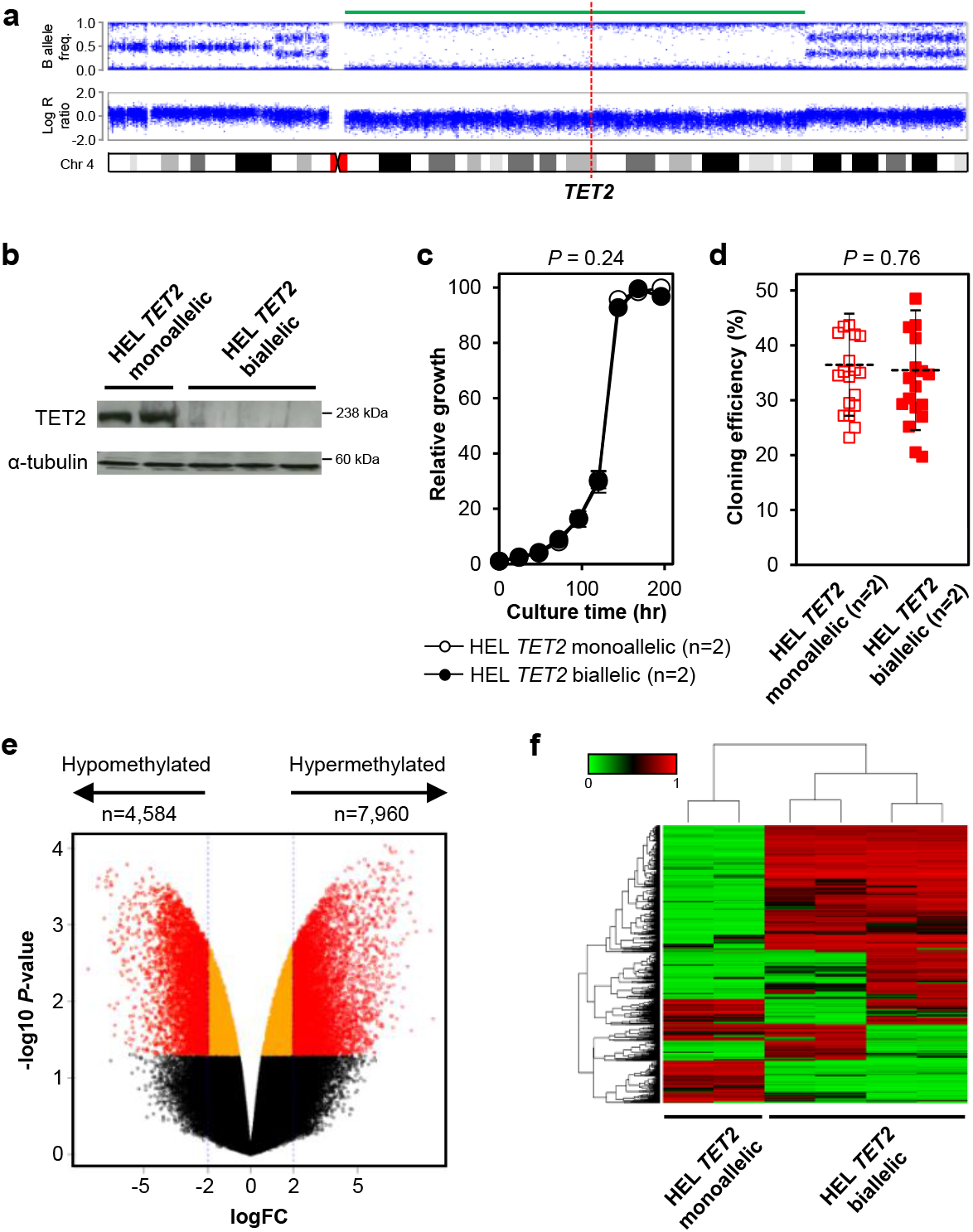
Complete loss of *TET2* expression confers a hypermethylation phenotype. (**a**) High density array copy number profile of chromosome 4 from HEL cells showing large deletion (green bar) affecting the q arm, including *TET2* (position indicated by dashed red line). (**b**) Immunoblot showing TET2 protein expression in two representative parental *TET2* monoallelic HEL cell clones (HEL *TET2* monoallelic) and three representative *TET2* CRISPR-Cas9-mutated HEL cell clones (HEL *TET2* biallelic). α-tubulin was used as loading control. (**c**) Growth kinetics in suspension culture of HEL *TET2* monoallelic (open circles) and HEL *TET2* biallelic (filled circles) cell clones. Cells were seeded at low density and growth (relative to initial density) was determined at regular intervals. Data represents mean and SD of indicated number of clones from three independent experiments. *P* value calculated by one-way ANOVA. (**d**) CE was calculated for HEL *TET2* monoallelic (open squares) and HEL *TET2* biallelic (filled squares) clones after 30 days culture in soft agar. Mean and SD of indicated number of clones from seven independent experiments are shown. *P* value calculated by 2-tailed Student’s t-test. (**e**) Volcano plot demonstrating differences in CpG methylation between HEL *TET2* monoallelic (n=2) and HEL *TET2* biallelic (n=4) clones. Plot was constructed using fold-change (Log_2_FC) values and adjusted *P*-values and points represent individual CpG probes, colored such that significantly differentially methylated probes (*P*<0.05 and |Log_2_FC|≥2) are in red. Orange points represent probes which reach significance (*P*<0.05) but are not differentially methylated (|Log_2_FC|<2) and black points represent non-significant (*P*≥0.05) probes. (**f**) Unsupervised hierarchical clustering of the top 1,500 differentially methylated CpG probes across all samples resulted in distinct clustering of parental HEL *TET2* monoallelic (n=2) and HEL *TET2* biallelic (n=4) cell clones. Rows in the heatmap represent CpG probes and vertical columns are cell clones. Color key indicates level of methylation at CpGs.

Analysis on human methylation (450K) arrays revealed the acquisition of an overall hypermethylation phenotype in *TET2* biallelic clones compared to *TET2* monoallelic clones (Figure 2E; Table S2). Unsupervised hierarchical clustering demonstrated clustering of HEL clones based on *TET2* mutation status (Figure 2F).

When treated with 5’-Aza *in vitro*, HEL *TET2* biallelic clones had significantly lower CE (*P*=0.003) and proliferation in liquid culture (*P*<0.001) compared to isogenic *TET2* monoallelic clones (Figure 3A-B). In contrast, biallelic *TET2* mutation did not affect CE or cell proliferation following treatment with Ara-C or daunorubicin (Figure 3A-B). Having demonstrated that HEL cells null for TET2 protein expression were sensitive to 5’-Aza we hypothesized that TET2 protein levels could affect sensitivity to 5’-Aza. In order to test this, we quantified TET2 protein levels in a panel of 10 AML cells lines and determined sensitivity to 5’-Aza (Figure 3C). There was significant correlation between TET2 protein levels and 5’-Aza IC90 (R^2^=0.77, *P*=0.0008) and IC50 (R^2^=0.88, *P*<0.0001) (Figure 3C). THP1 cells had the highest TET2 protein expression and were the most resistant to 5’-Aza. In contrast, SKM1 cells had the lowest TET2 protein expression and were the most sensitive to 5’-Aza. Intriguingly, SKM1 cells have a monoallelic *TET2* mutation (c.4253_4254insTT, p.1419fsX30)^9^ but are phenotypically null for TET2 protein expression despite having an intact WT *TET2* allele (Figure 3C). As such, unlike biallelic *TET2* deletion which renders cells null for TET2 expression, monoallelic *TET2* mutation is not necessarily a good indicator of TET2 protein expression.

**Figure 3.**
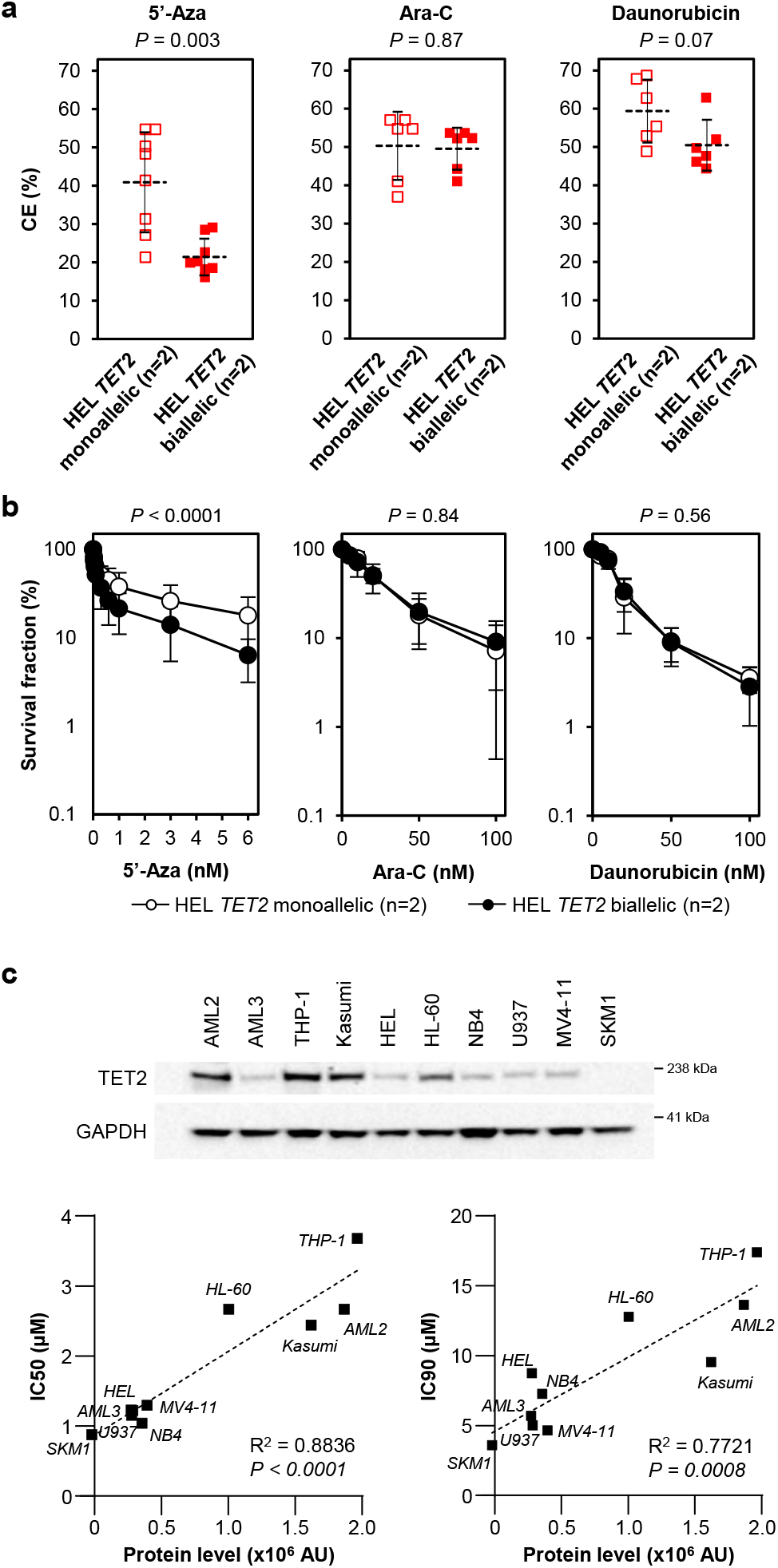
Cells with biallelic *TET2* mutation are sensitive to the hypomethylating agent, 5’-Aza in *in vitro* model systems. (**a**) Parental *TET2* monoallelic HEL cell clones (HEL *TET2* monoallelic; open squares) and *TET2* CRISPR-Cas9-mutated HEL cell clones (HEL *TET2* biallelic; filled squares) were cultured in soft agar supplemented with 1nM 5’-Aza (left), 20nM Ara-C (center) or 20nM daunorubicin (right) and CE (relative to respective vehicle control-treated cells) was determined after 30 days. Mean and SD of indicated number of clones from three independent experiments are shown. *P* values calculated by Student’s t-test (2-tailed). (**b**) Parental *TET2* monoallelic HEL cell clones (HEL *TET2* monoallelic; open circles) and *TET2* CRISPR-Cas9-mutated HEL cell clones (HEL *TET2* biallelic; filled circles) were treated with 5’-Aza (left), Ara-C (center) or daunorubicin (right) and cell density (relative to respective vehicle control-treated cells) was determined after 96 hrs. Data represents mean and SD of indicated number of clones from three independent experiments. *P* values calculated by two-way ANOVA. (**c**) Western blot (top panel) showing TET2 protein expression in a panel of 10 AML cell lines. GAPDH was used as a loading control. TET2 protein expression was quantified in each cell line and plotted against 5’-Aza IC50 (left) and IC90 (right) values. AU represents arbitrary units as measured by the Fuji LAS-300 Image Analyzer System.

We next determined whether biallelic *TET2* mutation sensitizes AML cells to 5’-Aza *in vivo* in an orthotopic xenograft mouse model. We used a competitive engraftment approach and an allele-specific qPCR assay for the WT and CRISPR-Cas9-modified *TET2* alleles to determine preferential engraftment and/or elimination of either cell clone. Specifically, following prior validation of the qPCR assay (Figure S6), HEL *TET2* monoallelic and HEL *TET2* biallelic cell clones were co-injected intrafemorally (IF) in a 1:1 ratio into *Rag2*^*−/−*^*Il2rg*^*−/−*^ mice (day 0; Figure 4A). Physical symptoms associated with the proliferation of AML cells became apparent approximately 4 weeks post-IF injection. On day 28 mice began once daily treatment with 5mg/kg 5’-Aza (or vehicle control (VC)) for a total of 5 days, and were then euthanized 3 days later for sample collection and qPCR analysis (Figure 4A). Human *TET2* DNA consistently amplified using DNA from injected femurs, as well as non-injected femurs, spleens and other organs showing evidence of AML infiltration (32 individual samples from VC-treated mice and 23 from 5’-Aza-treated mice). There was no overall mean preferential amplification of either the intact WT or CRISPR-modified *TET2* allele in all 32 tissue samples from VC-treated mice (Figure 4B), although the CRISPR-modified *TET2* allele slightly dominated in spleen samples (*P*=0.047; Figure S7) suggesting preferential engraftment of TET2 null cells specifically in this tissue. Conversely, in 5’-Aza-treated mice the WT *TET2* allele was dominant in 19 of 23 (83%) tissue samples (Figure 4B), demonstrating significant negative selection against TET2 null cells (*P*=0.0004) as a result of 5’-Aza treatment. There was also significant negative selection of TET2 null cells specifically in the BM of 5’-Aza-treated mice (*P*=0.014) with 2 femur samples completely negative for the CRISPR-modified *TET2* allele following 5’-Aza treatment (Figure 4B).

**Figure 4.**
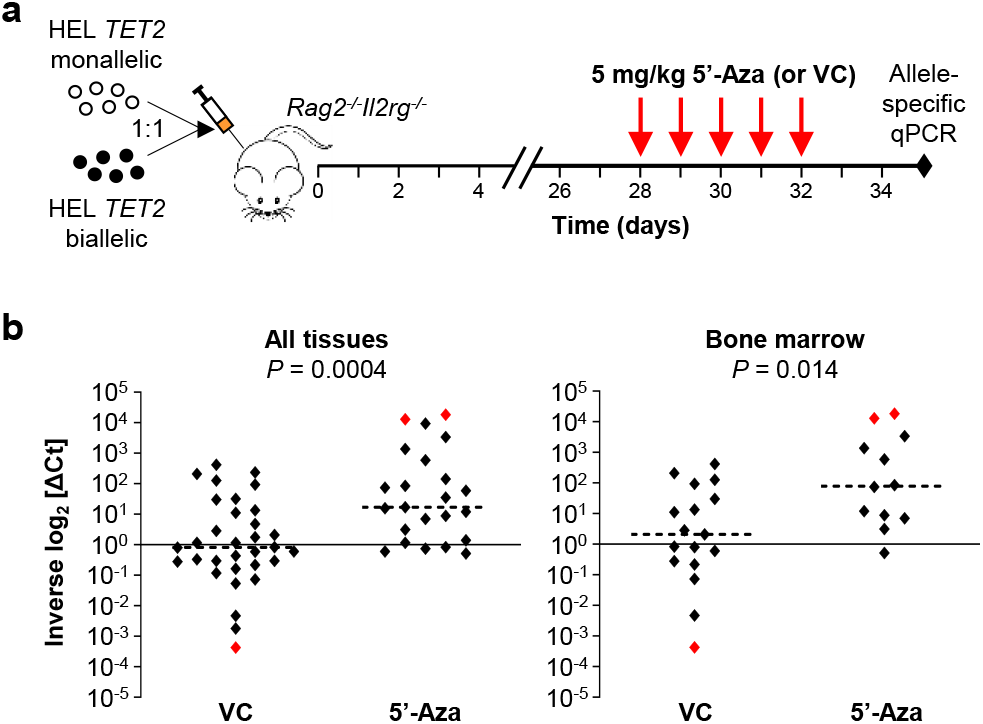
Cells with biallelic *TET2* mutation are subject to 5’-Aza-induced negative selection in an orthotopic AML mouse model. (**a**) Schematic of orthotopic AML mouse model. HEL *TET2* monoallelic and HEL *TET2* biallelic cell clones were co-injected in a 1:1 ratio into the femurs of *Rag2*^*−/−*^*Il2rg*^*−/−*^ mice. Treatment with 5-Aza (5mg/kg daily for 5 days) or VC was initiated on day 28 (post-injection) and tissues were harvested on day 35 for *TET2* allele-specific qPCR analysis. (**b**) Tissue samples collected from mice were analyzed by custom *TET2* allele-specific qPCR assay. Shown are inverse Log_2_ [ΔCt] values which represent relative expression of the WT versus the 4bp deleted *TET2* allele in individual samples, and are the means of triplicate reactions. Inverse Log_2_ [ΔCt] of 1 indicates a 1:1 ratio between the WT and 4bp deleted *TET2* alleles (and hence HEL *TET2* monoallelic and HEL *TET2* biallelic clones), whereas inverse Log_2_ [ΔCt] > 1 or inverse Log_2_ [ΔCt] < 1 indicates dominance of the WT (HEL *TET2* monoallelic) or 4bp deleted (HEL *TET2* biallelic) allele, respectively. Red points indicate samples which were dominated entirely by one cell clone. Horizontal dashed lines represent median inverse Log_2_ [ΔCt] values across all samples from VC- or 5’-Aza-treated mice. The left panel shows data from all harvested tissues (BM, PB, spleen and tumors) and the right panel shows data from BM only. *P* values comparing inverse Log_2_ [ΔCt] values from VC and 5’-Aza-treated mice were calculated using a Mann-Whitney test.

### Gene expression analysis identifies downregulation of small nuclear ribonucleoprotein complex components in cells with biallelic *TET2* mutation

RNA sequencing analysis was performed to identify differentially expressed genes and potential mechanisms responsible for 5’-Aza sensitivity in HEL *TET2* biallelic cell clones. HEL cell clones clustered broadly by *TET2* genotype (Figure 5A), suggesting that complete loss of TET2 protein significantly impacted on transcription. Differential expression analysis identified 695 significantly differentially expressed transcripts (*Padj*<0.05; |Log_2_FC|≥0.3) in HEL *TET2* biallelic clones compared to HEL *TET2* monoallelic clones (Figure 5B; Table S3) and gene ontology analysis identified several significantly affected cellular components (Table S4), of which the spliceosomal small nuclear ribonucleoprotein (snRNP) complex (GO:0097525) was the most significantly affected (*Padj*=8.7 × 10^−4^) (Figure 5C; Table S5). Likewise, spliceosomal tri-snRNP complex assembly (GO:0000244) was identified as the most significantly affected biological process (*Padj*=3.4 × 10^−4^; Table S6). Downregulation of protein expression in TET2 null cells was confirmed for LSM8, consistent with transcript expression data (Figure 5D). Significant differences in expression were also identified for other genes that could potentially affect cellular response to 5’-Aza (Table S3). These included *ABCB1* (*MDR1*)^10^, encoding a member of the ATP-binding cassette (ABC) family of drug transporters, which was down-regulated in TET2 null cells at the transcript (*Padj*=8.1 × 10^−4^) and protein level (Figure 5D).

**Figure 5.**
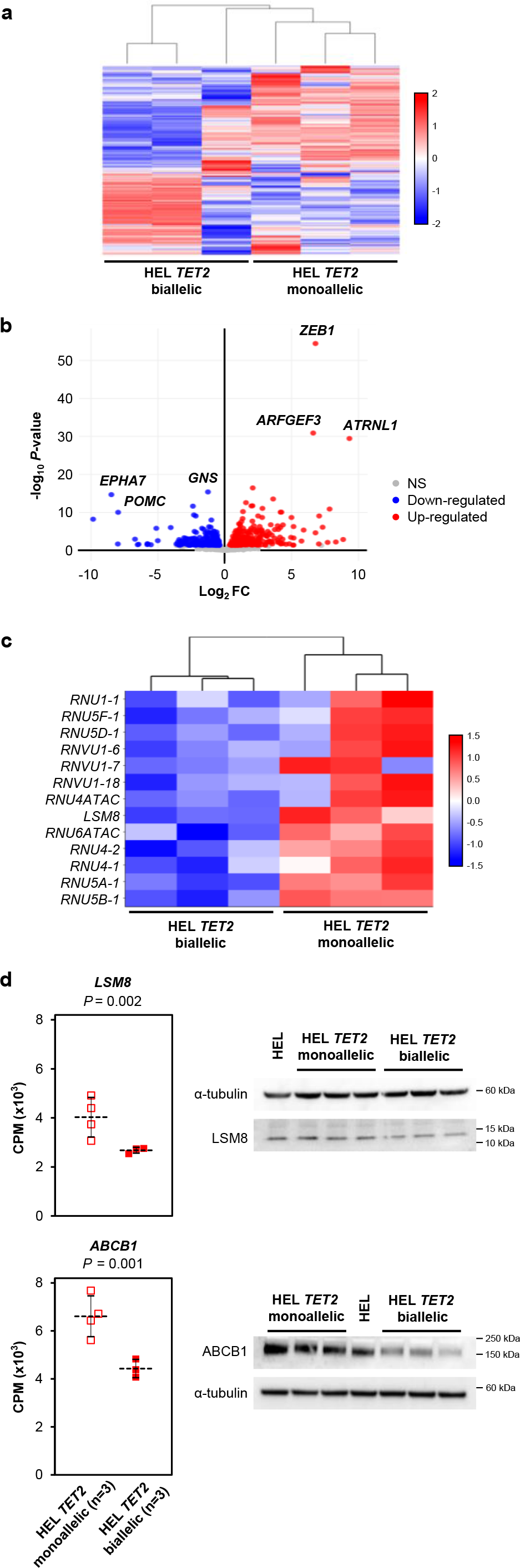
Differential gene expression in AML cells with monoallelic and biallelic *TET2* mutation. (**a**) Unsupervised hierarchical clustering of the top 1,500 differentially expressed transcripts in parental *TET2* monoallelic HEL cell clones (HEL *TET2* monoallelic; n=3) and TET2 CRISPR-Cas9-mutated HEL cell clones (HEL *TET2* biallelic; n=3). Horizontal rows of the heatmap represent individual transcripts and each vertical column is a cell clone. Color indicates relative expression with down-regulated and up-regulated transcripts indicated in blue and red, respectively. (**b**) Volcano plot demonstrating significant differential gene expression (*P*-value < 0.05 and |Log_2_FC| ≥ 0.3) in *TET2* CRISPR-Cas9-mutated HEL cell clones (HEL *TET2* biallelic; n=3) relative to parental HEL clones with monoallelic *TET2* mutation (HEL *TET2* monoallelic; n=3). Plot was constructed using fold-change (Log_2_ FC) values and adjusted *P*-values and points represent individual gene transcripts. Shown are 326 significantly down-regulated transcripts (blue) and 369 significantly up-regulated transcripts (red). Non-significant (NS) transcripts (*P*-value ≥ 0.05) are represented by grey points. Genes with particularly significant differential expression are labelled. (**c**) Heatmap showing differential expression of components of the spliceosomal snRNP complex (GO:009752) in parental *TET2* monoallelic HEL cell clones (HEL *TET2* monoallelic; n=3) and *TET2* CRISPR-Cas9-mutated HEL cell clones (HEL *TET2* biallelic; n=3). Horizontal rows represent genes and each vertical column is a cell clone. Color indicates relative expression, as in (a). (**d**) Transcript expression (expressed as counts per million (CPM) reads) of *LSM8* (top) and *ABCB1* (bottom) in parental *TET2* monoallelic HEL cell clones (HEL *TET2* monoallelic; open bars) and *TET2* CRISPR-Cas9-mutated HEL cell clones (HEL *TET2* biallelic; filled bars). Data represents the mean and SD of indicated number of clones. *P* values are from differential expression analysis. Western blots to the right of the charts show corresponding protein expression in the individual cell clones included in RNA sequencing analysis. α-tubulin was used as a loading control.

### Biallelic *TET2* alterations in AML patients with cytogenetically discernible chromosome 4 aberrations

The *TET2* locus can be somatically affected via numerous mechanisms, although how these manifest to give rise to biallelic mutation remains unclear. In order to investigate this we screened the SAL biobank for AML patients presenting with a cytogenetically discernible aberration affecting chromosome 4; a population likely to be enriched for *TET2* alterations. Ninety-two (6%) of 1560 recruited AML cases had a chromosome 4 aberration visible cytogenetically, and sufficient material for our analyses was available for 30 cases (Table S7). Gains affecting *TET2* were discernible by SNP array in 6 patients (all with trisomy 4 visible cytogenetically), which included two cases with homozygosity affecting most of the long arm of chromosome 4 (Figure 6A). One of these two patients (UPN25) also had a *TET2* base substitution (c.4133G>A, p.Cys1378Tyr) carried by almost 100% of the cells (Figure 6A and Figure S8) and therefore has biallelic *TET2* mutation. Six cases had loss of genetic material affecting *TET2* discernible from SNP array data, which included three cases (UPN09, UPN10 and UPN18) with large deletions and two cases (index case (UPN01) and UPN30) with a focal deletion in one allele and a nonsense mutation in the other allele (Figure 6A and Figure S8). The sixth case (UPN28) had trisomy 4 but with a focal 585Kb deletion affecting the entire *TET2* gene, resulting in copy number reduction (< 2 copies) and loss of heterozygosity (Figure 6A and Figure S8). A further 6 cases had copy number alterations (5 with loss, 1 with gain) on chromosome 4 which did not affect the *TET2* locus (Figure 6A). The remaining 12 cases had no evidence of *TET2* base substitution or gain/loss of material on chromosome 4. In summary, in a panel of 30 AML cases with a cytogenetically discernible chromosome 4 abnormality, 7 patients (23%) had loss of function *TET2* mutation, including 4 (13%) with monoallelic *TET2* mutation (all of which were whole gene deletions reducing *TET2* copy number) and 3 patients (10%) with biallelic *TET2* mutation (whole gene deletion plus nonsense mutation in two cases, and biallelic base substitutions resulting from a whole chromosome gain and loss of heterozygosity in the third case). These data demonstrate that *TET2* alterations are complex, involving gains or losses of material in combination with base substitution mutations.

**Figure 6.**
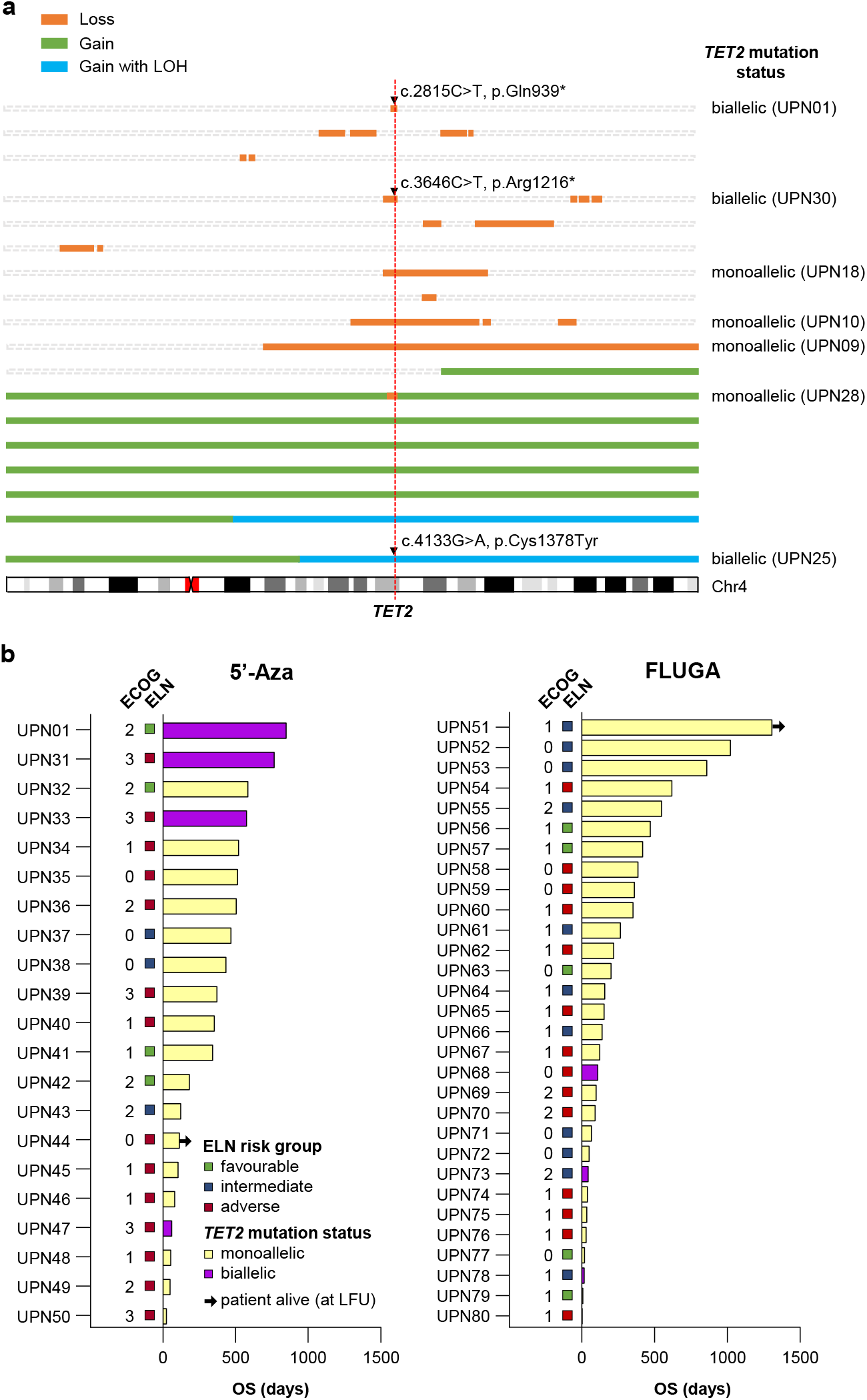
Somatic mutations affecting the *TET2* locus in AML patients with cytogenetic abnormality of chromosome 4 and response to treatment in AML patients with *TET2* mutation. **(a)** Illustrated are regions of copy number gain (green), gain with concomitant LOH (blue) and loss (orange) affecting chromosome 4 (discerned using high-density SNP array) in 18 AML patients with cytogenetically detectable abnormalities of chromosome 4. Base substitution mutations (indicated by black triangles) were determined by *TET2* exon sequencing. The vertical dashed red line indicates the location of the *TET2* gene. The mutation status of the 7 patients with loss of function *TET2* mutations are indicated to the right. Patient ID numbers are shown in parentheses for these patients. **(b)** Swimmer plots showing patients with *TET2* mutated AML treated with either 5’-Aza (left) or low-dose Ara-C plus fludarabine (FLUGA) (right). The AML index case (UPN01) is included in the 5’-Aza swimmer plot for reference. Patients with biallelic *TET2* mutation (UPN01, UPN31, UPN33, UPN47, UPN68, UPN73 and UPN78) are represented by purple bars. All other patients had monoallelic *TET2* mutation discerned by whole exome sequencing and are represented by pale yellow bars. European LeukemiaNet (ELN) favorable, intermediate and adverse risk groups are represented by green, blue and red squares, respectively. ECOG, Eastern Cooperative Oncology Group performance score.

### Biallelic *TET2* alterations in AML patients treated with 5-Aza

Having described a single patient with biallelic *TET2* mutation (UPN01) who responded very well to 5-Aza we sought to determine whether biallelic *TET2* mutation affected response in other patients treated with 5-Aza. AML patients over the age of 65 years were recruited to the PETHEMA FLUGAZA phase 3 clinical trial and were randomized to receive either 5-Aza or low-dose Ara-C plus fludarabine (FLUGA)^6^. Fifty patients had a *TET2* mutation identified by targeted sequencing which included 6 patients with a mutant allele frequency >85% indicative of biallelic *TET2* mutation (Table S8). None of the three patients with biallelic *TET2* mutation randomized to the FLUGA arm (UPN68, UPN73, UPN78) achieved CR and all had relatively short OS (111, 45 and 17 days) (Figure 6B; Table S8). In contrast, 2 of the 3 patients with biallelic *TET2* mutation randomized to the 5-Aza arm achieved CR (UPN31 and UPN33) and had prolonged OS (767 and 579 days) (Figure 6B; Table S8). The third patient treated with 5-Aza (UPN47) failed to achieve CR and died (day 62) after cycle 1 with progressive disease (Figure 6B; Table S8). All three patients with biallelic *TET2* mutation treated with 5-Aza had European LeukemiaNet (ELN) adverse risk AML and an Eastern Cooperative Oncology Group (ECOG) performance score of 3.

## Discussion

Up to 30% of AML patients present with a somatically acquired *TET2* mutation^11-17^, although those with biallelic mutation represent a small minority of all *TET2*-mutated AML cases^17-19^. The prognostic effect of *TET2* mutation in AML treated with anthracycline and nucleoside analogue-based regimens remains controversial^13,15,16^, although meta-analyses suggest an association with poor prognosis^20,21^. As such, there is an urgent clinical need to identify novel therapeutic approaches to improve outcome of *TET2*-mutated AML. Some studies have reported an association between *TET2* mutation and favorable outcome of myelodysplastic syndrome (MDS) following treatment with hypomethylating chemotherapy such as 5’-Aza^22-26^, although other studies have not replicated these findings^27^. Our data demonstrate that single-agent 5’-Aza treatment of AML harboring biallelic *TET2* mutation can give rise to long term CR, including in patients with disease refractory to standard remission induction chemotherapy and with adverse risk disease or poor performance status. Furthermore, using isogenic model systems, we demonstrate that biallelic *TET2* mutation confers cellular hypersensitivity to 5’-Aza *in vitro*, as well as significant negative selection when competitively xenografted with monoallelic *TET2*-mutated cells into BM of mice.

An effect of mutant *TET2* gene dosage on response to therapy is perhaps not surprising given the evidence demonstrating that mutant allele dose also affects disease development. Monoallelic (*Tet2*^+/-^) and biallelic *Tet2* deletions (*Tet2*^-/-^) both result in myeloid malignancy in animal models, but the latency and OS are significantly shorter in *Tet2* null animals^28,29^. Furthermore, *Tet2* null (*Tet2*^-/-^) mice with myeloid disease also have more pronounced splenomegaly compared to heterozygous (*Tet2*^+/-^) littermates^28^, and splenomegaly (and extramedullary disease) is a general feature of myeloid disease developing in *Tet2* knockout mouse models^29,30^. Consistent with this, young healthy mice null for *Tet2* have elevated extramedullary hematopoiesis in the spleen, which develops into splenomegaly concomitant with the onset of myeloid dysplasia^31^. These observations are consistent with our data demonstrating a significant competitive advantage of TET2 null human cells to populate the spleen of engrafted animals and also splenomegaly and extramedullary disease in the index patient (UPN01) reported herein. Taken together, these data suggest that TET2 loss could predispose to myeloid disease characterized by splenomegaly and extramedullary disease in general, which is mutant *TET2* gene dosage-dependent, although investigation in large patient cohorts is warranted.

Despite the observed negative selection against cells with biallelic *TET2* mutation *in vivo*, 5-Aza treatment rarely resulted in the complete elimination of TET2 null cells, consistent with data from the index patient in whom 5’-Aza-induced morphological remission was characterized by the persistence of cells with biallelic *TET2* mutation. Mutation persistence in morphological remission has been reported for several leukemia driver genes, including those characteristic of age-associated clonal hematopoiesis such as *TET2, DNMT3A, SRSF2, RUNX1* and *ASXL1*^32-35^. Likewise, persistence of *Tet2*-mutated cells has also been reported in animal models treated with 5’-Aza^36^. Our data demonstrate that cells with monoallelic and biallelic *TET2* mutation have significantly different genomic methylation profiles, and although we observed a genome-wide shift towards hypermethylation in cells with biallelic *TET2* mutation, the effect was relatively modest and there were also large numbers of CpG sites that became hypomethylated. Consistent with this, we also noted up-regulated transcript levels for numerous genes. As such, it seems unlikely that global genomic DNA methylation and concomitant global loss of expression is responsible for the observed sensitivity to 5’-Aza. Consistent with this model, pre-treatment DNA methylation levels do not correlate with clinical response in MDS to decitabine, the deoxynucleotide analogue of 5’-Aza^37^. Rather, the prevailing evidence suggests that the underlying mechanism conferring sensitivity to 5’-Aza is gene/pathway specific, and our investigations identified significant down-regulation of spliceosomal small nuclear ribonucleoprotein (snRNP) complex components in cells with biallelic *TET2* mutation. The snRNP pathway has previously been implicated as a determinant of cellular sensitivity to 5’-Aza^38^, although the underlying mechanisms remain to be fully deciphered.

Mutations in other genes operating in the TET2 hydroxymethylation pathway are also reported in AML, including *IDH1, IDH2* and *WT1*. Mutations in *IDH1* and *IDH2* inhibit TET2 function (and TET1 and TET3) via production of 2-hydroxyglutarate^39^, but are always heterozygous in AML^40,41^, suggesting a strong oncogenic gain of function phenotype where monoallelic mutation is sufficient to drive transformation with no selective pressure for loss of the second WT *IDH1* or *IDH2* allele. In contrast, both monoallelic and biallelic *WT1* mutations are reported in AML^42^ and drive leukemogenesis via inhibition of TET2 specifically. As such, loss of WT1 phenocopies loss of TET2 function and gives rise to a similar hypermethylation signature and a similar hematopoietic differentiation phenotype^43^. It will therefore be important to determine whether mutations in *WT1*, and particularly biallelic mutation, confers sensitivity to 5’-Aza. *TET2* mutations have also been reported in up to 28% of MDS and MPN^12,17,44^ and up to 50% of angioimmunoblastic T-cell lymphoma (AITL), where they are associated with poor response to anthracycline-based chemotherapy^45^. However, there is evidence of sensitivity to 5’-Aza in *TET2*-mutated AITL cases^46^, with prolonged CR reported in one case with double (presumed biallelic) mutation^45^.

Models for reliably predicting response to 5’-Aza in AML would be of clinical benefit. Our study suggests that *TET2* mutational profiling or TET2 protein expression analysis could potentially identify a subgroup of patients acutely sensitive to hypomethylating therapy, suggesting an alternative first line therapy for frail AML patients or salvage therapy for patients with chemoresistant disease. There is potential value in advocating *TET2* mutational or protein expression profiling in elderly patients with AML, where disease is more likely to have evolved from *TET2* clonal hematopoiesis and therefore likely to be enriched for AML with biallelic *TET2* mutation and null for expression^17^. Indeed, clinical studies in elderly AML have already documented excellent responses to 5’-Aza in some patients^47^, although the impact of TET2 status would need to be confirmed in prospective studies in all age groups. Likewise, there is a case to be made for implementing *TET2* mutational or expression profiling in AML patients with extramedullary disease, and particularly splenomegaly, given our data linking biallelic *TET2* mutation with colonization of the spleen in conjunction with data from mouse models showing a proclivity of *Tet2* mutation to drive extramedullary hematopoiesis and myeloid disease.

In summary, the prevailing evidence argues in favor of investigating mutant *TET2* allele dosage and TET2 protein expression as a determinant of sensitivity to 5’-Aza in large prospective studies of AML and other hematological conditions characterized by TET2 loss of function. However, comprehensive *TET2* mutational profiling that includes both sequence and copy number analysis would be required to identify patients with potentially complex alterations affecting the *TET2* locus. Furthermore, TET2 expression profiling could identify patients with disease that is null/low for protein expression despite the presence of at least one WT *TET2* allele, and who might also benefit from 5’-Aza treatment.

## Data Availability

Data is available from the corresponding authors on request.

## Acknowledgments

The authors wish to acknowledge Professor J. Kotzerke for providing PET-CT images (Figure. S1). This work was funded by a specialist programme grant from Blood Cancer UK (#13044 to JMA). This work was also supported in part by a grant from the German Consortium for Translational Cancer Research (DKTK) Dresden (to FS), the J.G.W. Patterson Foundation (#30015.088.085/PA/IXS to JMA) and the Accelerator award CRUK/AIRC/AECC joint funder partnership.

## Authorship

### Contribution

FS, SEF and W-YL designed experiments, generated data, analyzed data and wrote the manuscript. HB, CE, BM, LR, DK, CD, DA, DN, RP, E-NS, CP, MF, TR, AA, MW, HA, CR, LW, GLJ, TM, GHJ, HJM, JF, KO, MM, OH, TH, SV, BA, RAD, FP, PM, JM-L and MB generated/collated data and/or advised on data analysis. JMA designed experiments, generated data, analyzed data, directed the research and wrote the manuscript. FS and JMA also conceived of the project and secured funding. All authors contributed to the final version of the manuscript.

### Conflict-of-interest disclosures

MM is employed by the Munich Leukemia Laboratory and TH is part owner of the Munich Leukemia Laboratory. The remaining authors declare no competing financial interests or conflicts of interest.

### Correspondence

James M Allan, Newcastle Centre for Cancer, Newcastle University, Newcastle Upon Tyne, UK; phone: +44 (0)191 208 4435; fax: +44 (0)191 208 4301. Friedrich Stölzel, Medizinische Klinik und Poliklinik I, Universitätsklinikum Dresden, TU Dresden, Dresden, Germany;

## Notes

### Clinical Trial

NCT02319135

### Author Declarations

AML patients with an abnormal chromosome 4 (UPN01-UPN30) were from the Study Alliance Leukemia (SAL) AML registry biobank in Dresden (Germany) which was approved by the Institutional Review Board of the Medical Faculty of Dresden (Ethikkomission der TU Dresden, Medizinische Fakultat Dresden; IRB #EK98032010). TET2 mutant patients over 65 years of age with newly diagnosed TET2 mutant AML (UPN31-80) were from the Programa para el Estudio de la Terapeutica en Hemopatias Malignas (PETHEMA) phase 3 FLUGAZA clinical trial (NCT02319135)6, which was approved by the Institutional Review Board of Hospital Le Fe, Valencia, Spain (Eudract=2014-000319-15). Written informed consent was received from all study participants.

