## Supplemental Material for "Biallelic *TET2* mutation sensitizes to 5’-azacitidine in acute myeloid leukemia"

SUPPLEMENTAL MATERIALS

Supplemental Methods

Figure. S1. Disease infiltration in the AML index case.

Figure. S2. Sanger sequencing of *TET2* exon 3 in the index AML case.

Figure. S3. Sanger sequencing of *NPM1* exon 11 in the index AML case.

Figure. S4. Sanger sequencing of *FLT3* exon 14 in the index AML case.

Figure. S5. Sanger sequencing of *TET2* exon 6 in HEL AML cells.

Figure. S6. Validation of qPCR assay for detection of WT and CRISPR-Cas9-mutated *TET2* alleles.

Figure. S7. Preferential engraftment of TET2-null cells in the spleen of *Rag2^−/−^ Il2rg^−/−^* mice.

Figure. S8. High density SNP array copy number profile of chromosome 4 from leukemic blast cells of AML patients UPN25, UPN28 and UPN30.

Table S1. Karyotypes of cell lines used in the study.

Table S2. Significantly differentially methylated CpGs (*P* < 0.05) in HEL cell clones with biallelic *TET2* mutations compared to parental clones with monoallelic *TET2* mutation.

Table S3. Significantly differentially expressed genes (*P* < 0.05 and |Log_2_FC| ≥ 0.3) in HEL cell clones with biallelic *TET2* mutations compared to parental clones with monoallelic *TET2* mutation.

Table S4. Gene ontology component analysis of significantly differentially expressed genes in HEL cell clones with biallelic *TET2* mutations compared to parental clones with monoallelic *TET2* mutation.

Table S5. Significantly differential expression (*P* < 0.05) of components of the snRNP complex (GO:0097525) in HEL cell clones with biallelic *TET2* mutations compared to parental clones with monoallelic *TET2* mutation.

Table S6. Gene ontology biological pathway analysis of significantly differentially expressed genes in HEL cell clones with biallelic *TET2* mutations compared to parental clones with monoallelic *TET2* mutation.

Table S7. Clinical characteristics and demographics of AML patients with a cytogenetically discernible chromosome 4 aberration (from SAL AML Biobank).

Table S8. Clinical characteristics and demographics of AML patients with *TET2* mutation enrolled in the PETHEME FLUGAZA phase 3 clinical trial.

Table S9. Primer sequences and PCR reaction conditions for Sanger sequencing of genes of interest.

Supplemental References

**Supplemental Methods**

**BM morphological assessment of the AML index case (UPN01)**

For morphological analyses at AML presentation and during follow up, smears were prepared from BM aspirates, stained with Giemsa and visualized according to routine diagnostic protocols.

**Cytogenetic analyses of UPN01**

G-banding analysis of metaphase chromosomes from short-term cultures established from presentation BM aspirate was performed using well-established techniques. Interphase FISH was performed using the XL *TET2* kit (Metasystems, Germany). Spectral karyotyping (SKY) was performed using the SKYPaint probe mixture kit (Applied Spectral Imaging, Israel) according to the manufacturer’s protocol, with the exception that the hybridization time was extended from two to three days.

**Nucleic acid preparation**

DNA was extracted from cultured cells, BM mononuclear cells (BMMNCs), peripheral blood (PB) or methanol:acetic acid-fixed cells using an appropriate Qiagen kit (Qiagen, Manchester, UK) or from saliva using an Oragene kit (DNA Genotek, Ottawa, Canada).

**SNP array genotyping**

SNP array genotyping was performed on DNA from BMMNCs using the OmniExpressExome (v1.4) platform and analyzed using GenomeStudio 2.0.3 (Illumina, San Diego, CA) with genotype, minor (‘B’) allele frequency (B / A + B) and logR ratio at each locus calculated using standard parameters (GenCall Threshold 0.15). SNP coordinates are based on human genome build 37. Regions of copy number loss were identified manually based on interrogation of LogR ratios and B allele frequencies.

**Whole exome sequencing**

Exome capture (using Agilent SureSelect Protocol v1.2), library preparation and sequencing of pooled DNA samples (from PB or saliva) was carried out by Oxford Gene Technology (Oxfordshire, UK) on the Illumina HiSeq2000 platform. Reads were mapped to human genome build 37 (hg19) using the Burrows-Wheeler Aligner MEM package^1^ and local realignment of mapped reads around potential insertion/deletion (indel) sites was carried out using Genome Analysis Toolkit^2^ (GATK; v1.6). Duplicate reads were marked using Picard (v1.98) and excluded from analysis. SNPs and indels were called using GATK HaplotypeCaller, with SNP novelty determined against dbSNP release 135. Variants were annotated with gene data from Ensembl. A read depth of at least 20x was achieved for a minimum of 95.61% of on-target regions.

**RNA sequencing and differential gene expression analysis**

Total RNA was extracted using the RNeasy micro kit (Qiagen) and quantified using a Qubit 2.0 Fluorometer with Qubit RNA BR assay kit (Thermo Fisher Scientific, MA, USA). Quality control, library preparation and sequencing on the NextSeq 550 platform (Illumina) was performed by Edinburgh Clinical Research Facility (Edinburgh, UK).

Sequencing reads were mapped to human genome build 37 (hg19) and annotated using STAR aligner^3^. Aligned reads were summarized over gene features using the Rsubread package^4^ (using featureCounts function) in R (v3.5.1). Read counts were normalized by expressing as CPM. Gene level differential expression analysis was performed on normalized read counts using DESeq2 (version 1.16.1)^5^. Resulting *P*-values were adjusted to control for the false discovery rate (FDR; 5%)^6^ and significantly differentially expressed genes were defined as those with FDR-adjusted *P* value < 0.05 and |Log_2_FC| ≥ 0.3.

**Illumina 450k arrays and differential methylation analysis**

DNA was extracted from HEL cell clones using a DNA Mini Kit (Qiagen) and sent for processing and hybridization to Infinium® HumanMethylation450 Beadchips (Illumina) by Eurofins Genomics (Galten, Denmark). Data processing and analysis was performed according to an established workflow^7^. Specifically, raw intensity data (IDAT) files containing methylated (M) and unmethylated (U) intensity measurements were imported into R and the minfi Bioconductor package^8^ was used to calculate detection p-values (*detP*), normalize data (using the preprocessFunnorm function) and generate β (β = M/(M+U+100)) and M (M = log_2_(M/U)) values for individual CpG probes. Poorly performing probes (*detP* < 0.01) and those interrogating SNPs were removed, leaving 410,811 probes in the final dataset. The limma Bioconductor package^9^ was used to identify significantly differentially methylated probes based on *TET2* mutation status (monoallelic vs biallelic) using M values. Resulting *P*-values were adjusted to control for FDR (5%)^6^ and significantly differentially methylated CpGs were defined as those with FDR-adjusted *P*-value < 0.05 and |Log_2_FC| ≥ 2. Unsupervised hierarchical clustering based on M values was performed in R with scaling by standard deviation.

***TET2*, *NPM1* and *FLT3* mutation analysis**

Whole gene *TET2* mutation analysis of the SAL abnormal chromosome 4 cases was performed by the MLL Munich Leukemia Laboratory (Munich, Germany) on DNA from fixed BMMNCs via the generation of 27 exon-specific amplicons using the FastStart High Fidelity PCR system kit (Roche Applied Science, Penzberg, Germany) as previously described^10,11^. Mutation of *TET2* exon 3 in the index AML case (UPN01), as well as *NPM1* exon 11 and *FLT3* exon 14, was confirmed by Sanger sequencing. PCR reactions consisted of 0.5 units ThermoPrime Taq DNA polymerase with 1x ReddyMix PCR buffer (Thermo Fisher Scientific), 1.5mM MgCl_2_, 10pmol primers, 0.2mM (each) dNTPs (Invitrogen Life Technologies, Paisley, UK) and 100ng template DNA in a total volume of 20μl. Primer sequences and thermal cycling conditions for individual amplicons are shown in Table S9. PCR products were purified using the QIAquick® PCR Purification kit (Qiagen) and sequenced using the indicated primers (Table S9) by Source BioScience (Nottingham, UK). Mutation in *TET2* was determined on the PETHEMA-FLUGAZA AML clinical trial patients from whole exome sequencing, as previously described^12^.

**Western Immunoblotting**

Cellular proteins were extracted using Phosphosafe reagent (Millipore Ltd, Watford, UK) and quantified by Pierce BCA assay (Thermo Fisher Scientific). Proteins were separated using Novex® NUPAGE 3-8% tris-acetate gels (Invitrogen Life Technologies), transferred to nitrocellulose membranes and immunoblotted according to routine techniques. Antibodies used were TET2 (Mab-179-050; Diagenode, NJ, USA), ABCB1 (G-1; Santa Cruz Biotechnology, Dallas, Texas), LSM8 (F-8; Santa Cruz Biotechnology), α-tubulin (T9026; Sigma-Aldrich) and GAPDH (0411; Santa Cruz). Appropriate HRP-conjugated secondary antibodies were from Agilent Technologies (CA, USA). Protein quantification was performed on immunoblots using the Fuji LAS-300 Image Analyser System (Raytek, Sheffield UK).

**Cell proliferation, drug sensitivity and cloning efficiency assays**

Cytotoxic agents were purchased from Sigma-Aldrich. Ara-C was reconstituted in DMSO and daunorubicin or 5-Aza in dH_2_O and aliquots were prepared and stored at -80°C. Stocks were diluted in CM immediately prior to use in cytotoxicity assays.

To compare cell proliferation between parental and CRISPR-Cas9-mutated HEL clones, exponentially growing cells were seeded at low density (2x10^4^ cells ml^-1^) in CM and counted using a hemocytometer at regular intervals up to 192 hours post-seeding. Cell growth at each timepoint was calculated relative to initial seeding density. Two-way ANOVA was used to test for significant differences in relative cell growth based on *TET2* mutation status.

For drug sensitivity experiments, cells were incubated in CM supplemented with appropriate concentrations of cytotoxic agent (5’-Aza, daunorubicin or Ara-C) or relevant vehicle control (VC) for 96 hours, after which viable cells were identified by trypan blue dye exclusion and counted using a hemocytometer. Survival fractions were determined at each drug concentration relative to VC-treated controls. Two-way ANOVA was used to test for significant differences in drug sensitivity based on *TET2* mutation status. Inhibition of proliferation in drug-treated cultures was compared to VC-treated cultures and used to calculate the IC50 and IC90 values in GraphPad Prism (PRISM 6.0.2, Graphpad Software).

For determination of CE, exponentially growing cells were seeded in soft agar [CM supplemented with 0.2% agarose] supplemented with cytotoxic agent (5’-Aza, daunorubicin or Ara-C) or VC. Macroscopically visible colonies were counted on day 30 and CE was calculated relative to number of cells initially seeded. Student’s t-tests (2-tailed) were used to identify significant differences in CE based on *TET2* mutation status.

All assays were performed in triplicate at a minimum and means ± SD were calculated.

**Figure. S1**

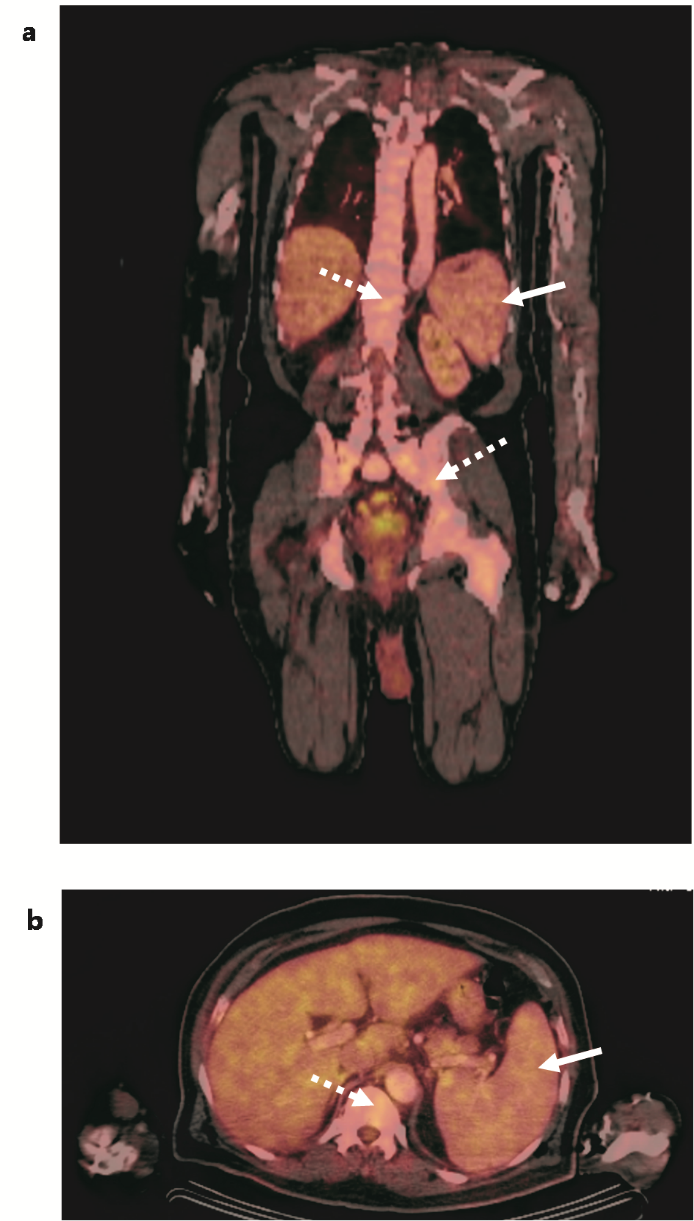

**Figure. S1. Disease infiltration in the AML index case.**

Coronal (**a**) and transversal (**b**) fused multiplanar reconstruction of ^18^Fluorodesoxy-Glucose Positron Emission Tomography – Computed Tomography imaging of the index AML patient (UPN01) at diagnosis, prior to induction chemotherapy. Splenomegaly (measuring 16.5×11×5.5 cm) is indicated by solid arrows. Enhanced hypermetabolic and focal uptake in the vertebrae and pelvic bone (dotted arrows) corresponds to the AML diagnosed in bone marrow and peripheral blood of the patient. Images courtesy of Professor J. Kotzerke.

**Figure. S2**

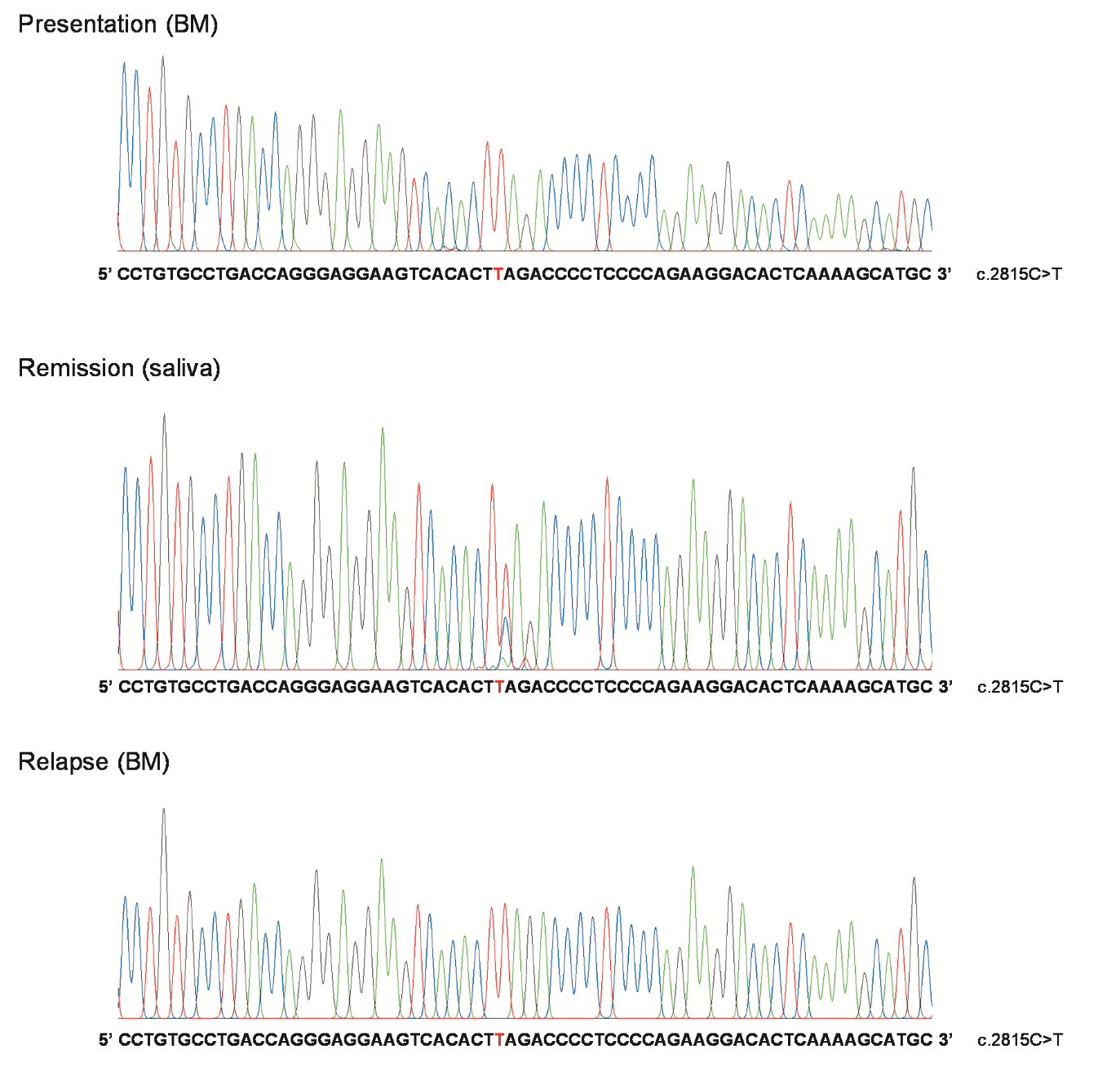

**Figure. S2. Sanger sequencing of *TET2* exon 3 in the index AML case.**

Sanger sequencing results for index AML patient at presentation (top), remission (middle) and relapse (bottom). A 65 bp section of forward read of *TET2* exon 3 surrounding the mutated base is shown in each case. The mutated base is shown in red. BM, bone marrow.

**Figure. S3**

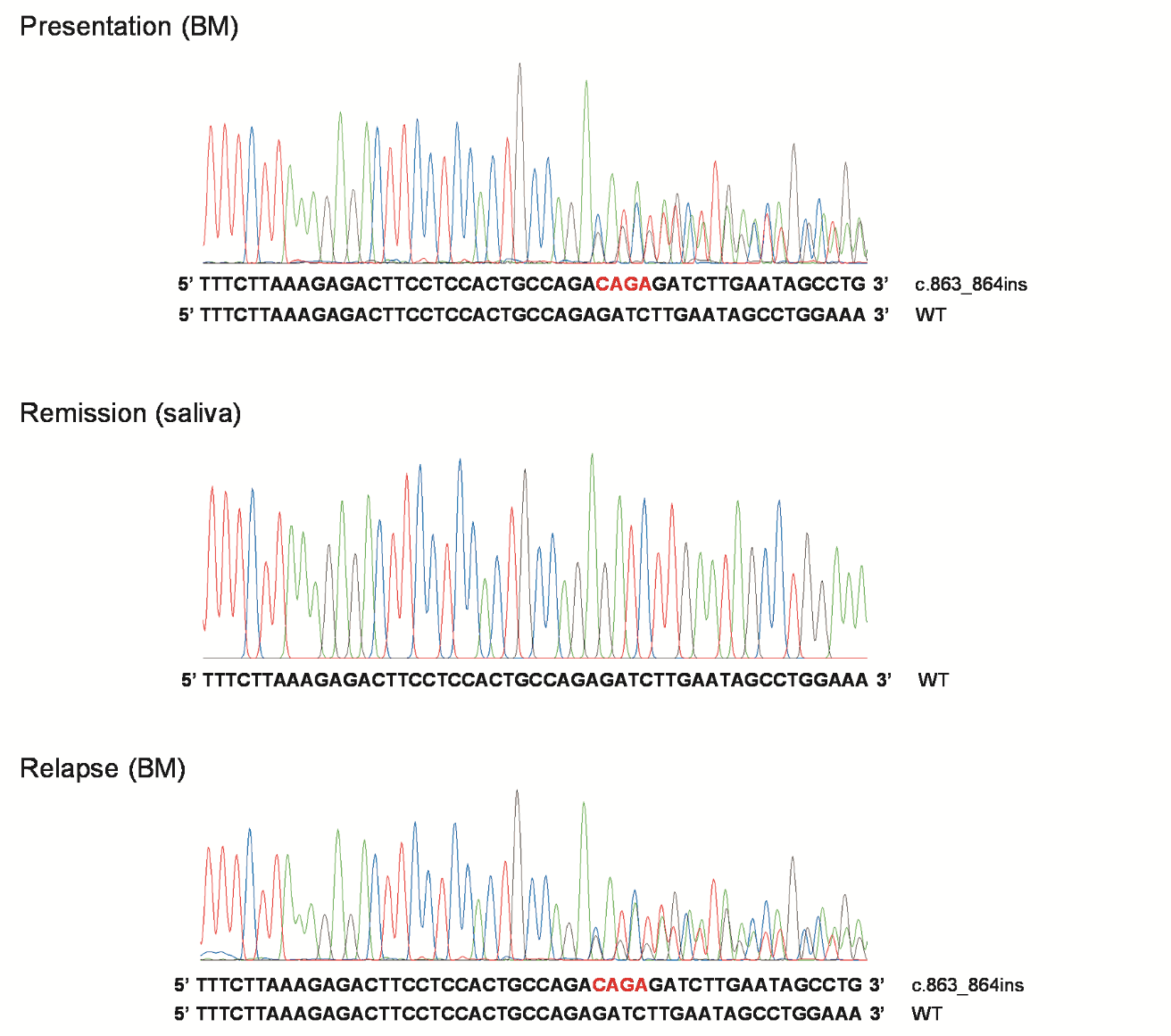

**Figure. S3. Sanger sequencing of *NPM1* exon 11 in the index AML case.**

Sanger sequencing results for index AML patient at presentation (top), remission (middle) and relapse (bottom). A 50 bp section of reverse complement read of *NPM1* exon 11 surrounding the insertion site is shown in each case. The 4 bp inserted sequence is shown in red. BM, bone marrow.

**Figure. S4**

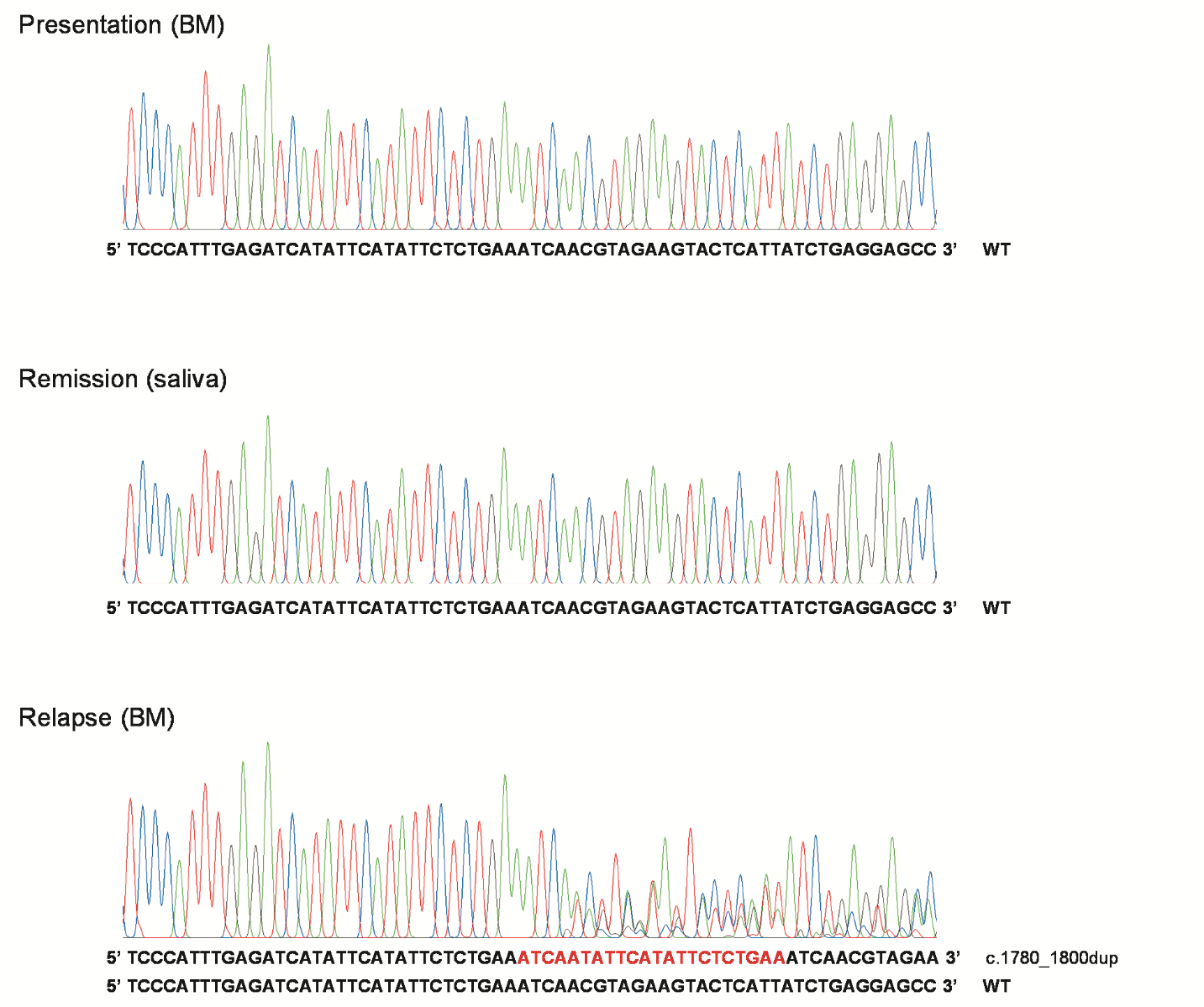

**Figure. S4. Sanger sequencing of *FLT3* exon 14 in the index AML case.**

Sanger sequencing results for index AML patient at presentation (top), remission (middle) and relapse (bottom). A 65 bp section of reverse complement read of *FLT3* exon 14 surrounding the duplication is shown in each case. The 21 bp duplicated sequence is shown in red. BM, bone marrow.

**Figure. S5**

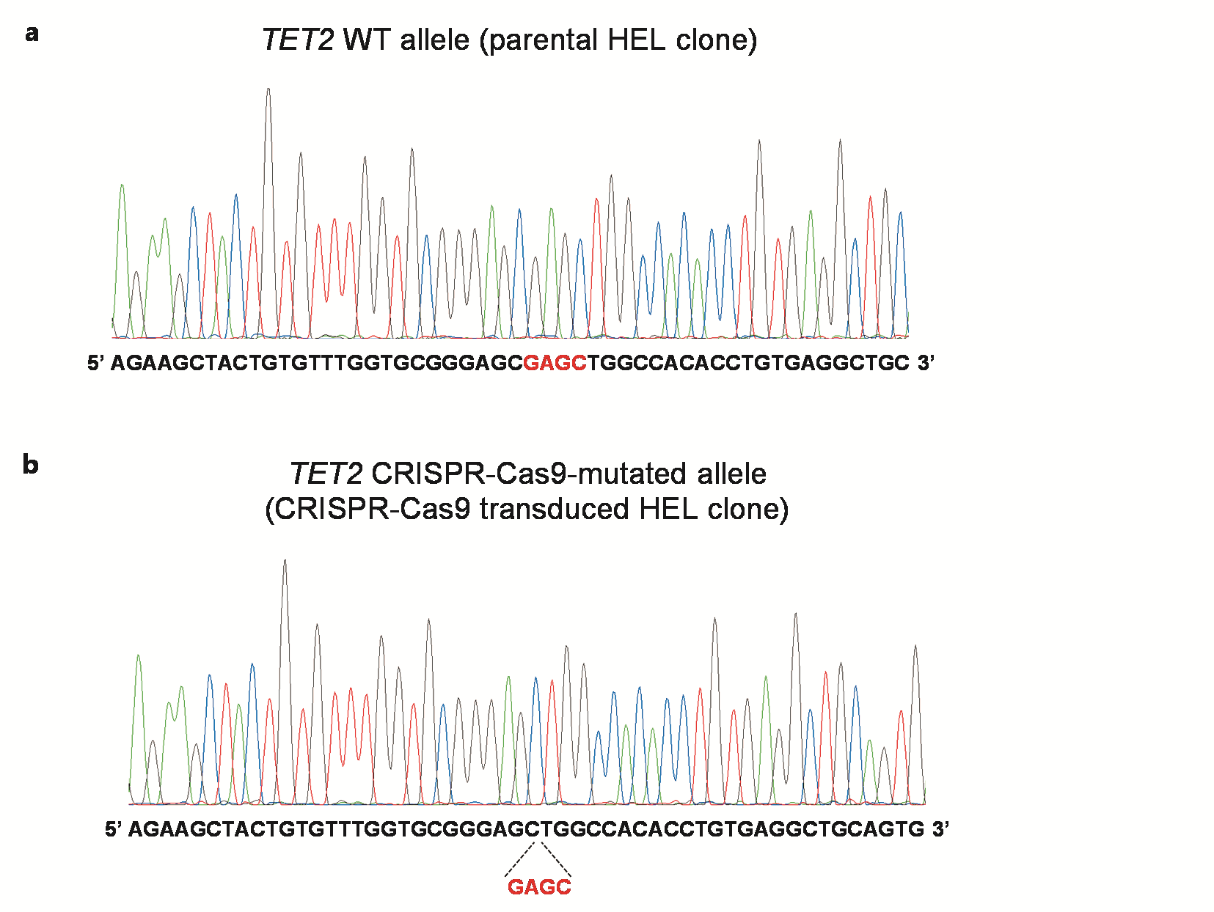

**Figure. S5. Sanger sequencing of *TET2* exon 6 in HEL AML cells.**

(**a**) Sanger sequencing of *TET2* exon 6 in a representative parental *TET2* HEL cell clone with WT sequence. (**b**) Sanger sequencing of *TET2* exon 6 in a *TET2* CRISPR-Cas9 transduced HEL cell clone with a 4bp deletion (highlighted in red).

**Figure. S6**

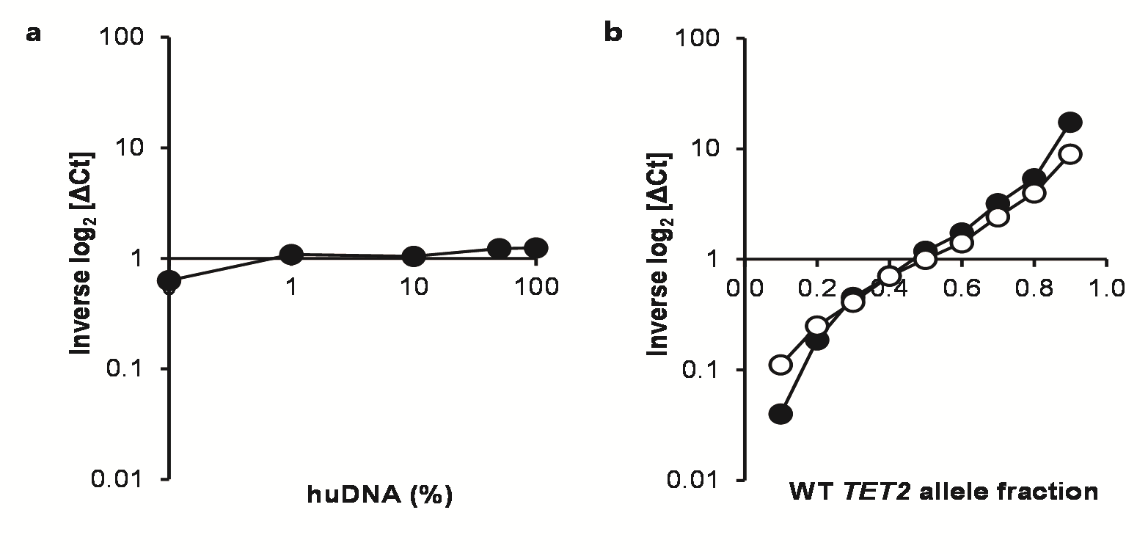

**Figure. S6. Validation of qPCR assay for detection of WT and CRISPR-Cas9-mutated *TET2* alleles.**

(**a**) Using a custom designed Taqman SNP Genotyping assay (see Methods), human DNA (huDNA) (from an equal mix of HEL *TET2* monoallelic and HEL *TET2* biallelic cells) was amplified in the presence of murine DNA in varying proportions and relative expression of the WT versus the mutated *TET2* allele (expressed as inverse Log_2_ [ΔCt]) was calculated. Equal amplification of both alleles is represented by a value of 1, whereas dominance of the WT or mutant allele is represented by inverse Log_2_ [ΔCt] values >1 or <1, respectively. As shown, amplification of huDNA was robust even when present at only 0.1% in murine DNA. Furthermore, no significant amplification bias of either WT or CRISPR-Cas9-mutated *TET2* alleles was observed. Amplification of either allele did not occur when only murine DNA was present in the reaction, confirming the assay to be specific for huDNA. Data represents the mean of triplicate reactions. (**b**) HEL *TET2* monoallelic and HEL *TET2* biallelic cells were mixed in varying ratios and extracted DNA was amplified using the custom Taqman SNP Genotyping assay as above. Closed circles demonstrate the actual experimentally-derived data (shown are means of triplicate reactions) and open circles represent the hypothetical data, assuming perfect amplification of both alleles at the designated ratios. As shown, there was no significant allele amplification bias when clones were mixed at ratios between 1:4 (WT allele fraction = 0.2) and 4:1 (WT allele fraction = 0.8). However, at ratios of 1:9 (WT allele fraction = 0.1) and 9:1 (WT allele fraction = 0.9) there was modest amplification bias in favour of the more abundant allele, presumably due to allelic drop-out of the less abundant allele.

**Figure. S7**

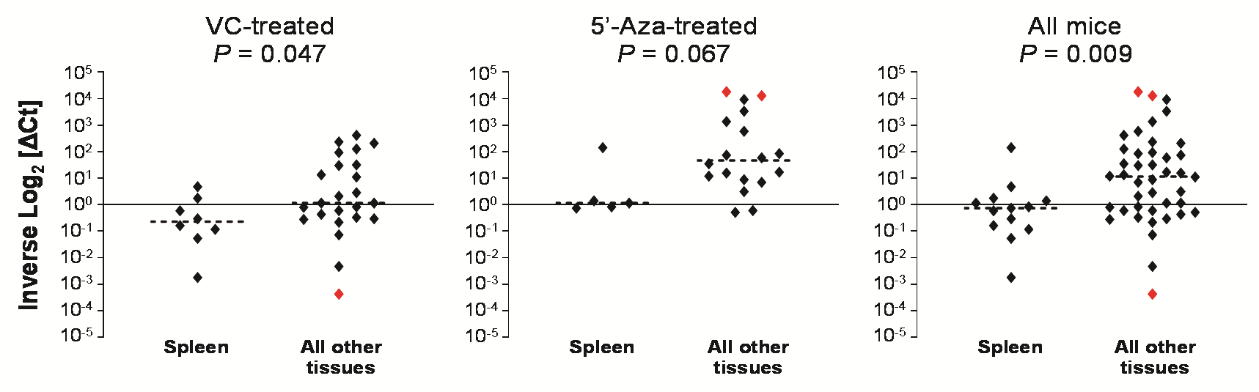

**Figure. S7. Preferential engraftment of *TET2*-null cells in the spleen of *Rag2^−/−^Il2rg^−/−^* mice.**

HEL *TET2* monoallelic and HEL *TET2* biallelic cell clones were co-injected in a 1:1 ratio into the femurs of *Rag2^−/−^Il2rg^−/−^* mice. Treatment with 5-Aza (5mg/kg daily for 5 days) or VC was initiated on day 28 (post-injection) and tissues were harvested on day 35 for analysis by custom *TET2* allele-specific qPCR assay. Shown are inverse Log_2_ [ΔCt] values which represent relative expression of the WT versus the 4bp deleted *TET2* allele in individual samples and are the means of triplicate reactions. Inverse Log_2_ [ΔCt] of 1 indicates a 1:1 ratio between the WT and 4bp deleted *TET2* alleles (and hence HEL *TET2* monoallelic and HEL *TET2* biallelic clones), whereas inverse Log_2_ [ΔCt] > 1 or inverse Log_2_ [ΔCt] < 1 indicates dominance of the WT (HEL *TET2* monoallelic) or 4bp deleted (HEL *TET2* biallelic) allele, respectively. Red points indicate samples which were dominated entirely by one cell clone. Horizontal dashed lines represent median inverse Log_2_ [ΔCt] values. *P* values comparing inverse Log_2_ [ΔCt] values from spleens versus all other tissues (BM, PB and tumours) in VC-treated (left), 5’-Aza-treated (center) or all mice regardless of treatment (right) were calculated using the Mann-Whitney test.

**Figure. S8**

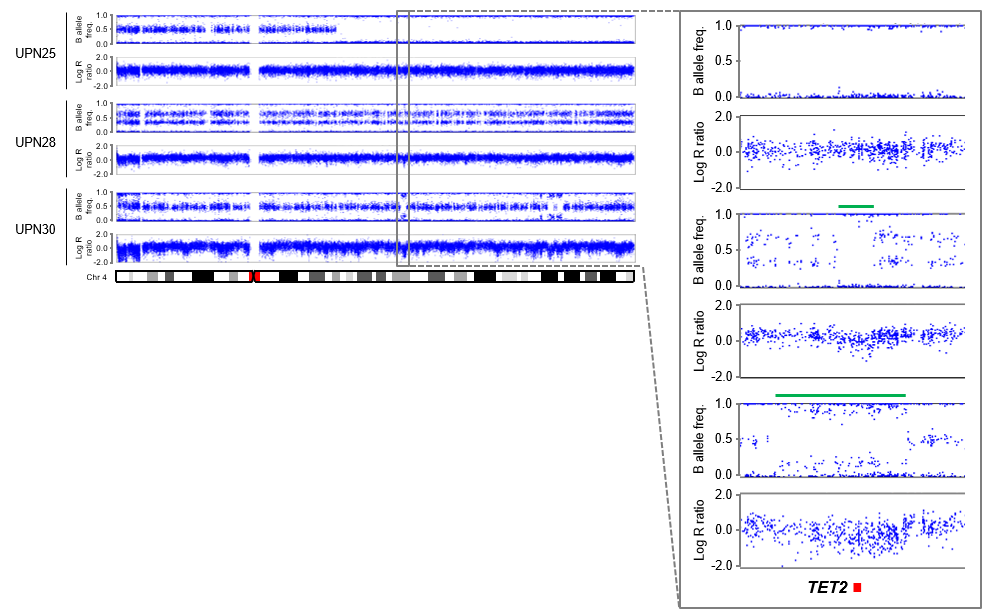

**Figure. S8. High density SNP array copy number profile of chromosome 4 from leukemic blast cells of AML patients UPN25, UPN28 and UPN30.**

Points represent individual SNPs which are aligned relative to their position on chromosome 4 (indicated by the ideogram below the plots). Copy number is measured as Log R ratio, with a value of 0 indicating a diploid SNP and positive and negative values indicating copy number gain and loss, respectively. B allele frequency represents the ratio of the two alleles of each SNP such that a value of 0.5 indicates allele heterozygosity and values of 0 and 1 indicate homozygosity. Inset shows expanded view of the region indicated by the grey box. Green bars highlight focal deletions within 4q24 in two of the patients. The location of *TET2* is shown below the plots.

**Table S1. Karyotypes of cell lines used in the study.**

| **Cell line** | **Karyotype** |
| --- | --- |
| AML-2 | 48(43-49)<2n>XY, +6, +8, der(1)inv(1)(p36q31)t(1;6) (q13;p12), der(2)t(2;17)(p23;q24.1)del(2)(q14.2q36), der(3)t(1;3)(p36;p25), ins(3;2)(q21;q14.2q36), t(5;8)(q11.2;q24), der(6)t(1;6)(q31;p12)t(3;6) (q26;q24), inv(12)(p13.3q13.2), t(13;14)(q32/33;q24.2), der(17)t(2;17)(p23;q24.1)^a^ |
| AML-3 | 48(45-50)<2n>X/XY, +1, +5, +8, der(1)t(1;18)(p11;q11), i(5p), del(13)(q13q21), dup(17)(q21q25)^a^ |
| THP-1 | 94(88-96)<4n>XY/XXY, -Y, +1, +3, +6, +6, -8, -13, -19, -22, -22, +2mar, add(1)(p11), del(1)(q42.2), i(2q), del(6)(p21)x2-4, i(7p), der(9)t(9;11)(p22;q23)i(9)(p10)x2, der(11)t(9;11)(p22;q23)x2, add(12)(q24)x1-2, der(13)t(8;13)(p11;p12), add(?18)(q21)^a^ |
| Kasumi-1 | 45<2n>X, -Y, -9, -13, -16, +3mar, t(8;21)(q22;q22), der(9)t(9;?)(p22;?), der(15)t(?9;15)((?q11;?p11)^a^ |
| HEL | 63(60-64)<3n>XYY, -2, -9, -10, -10, -11, -14, -16, -16, -17, -19, +20, +21, +2mar, del(2)(q32), t(3;6)(p13;q16), der(5)t(5;17)(q10;q10), der(6)t(1;6)(p13;p21), der(7)add(7)(p14;q32), add(8)(p21), der(9)t(9;?)(?;11)(p24;?)(?;q13), del(11)(q13), add(15)(p11), del(20)(q13), r(20)(p11q11), dup(21)(q11q22.3-qter), psu dic(22;9)t(9;?)(?;22)(p24;?)(?;p11-13)^a^ |
| HL-60 | 82-88<4n>XX, -X, -X, -8, -8, -16, -17, -17, +18, +22, +2mar, ins(1;8)(p?31;q24hsr)x2, der(5)t(5;17)(q11;q11)x2, add(6)(q27)x2, der(9)del(9)(p13)t(9;14)(q?22;q?22)x2, der(14)t(9;14)(q?22;q?22)x2, der(16)t(16;17)(q22;q22)x1-2, add(18)(q21)^a^ |
| NB4 | 78(71-81)<3n>XX, -X, +2, +6, +7, +7, +11, +12, +13, +14, +17, -19, +20, +4mar, der(8)t(8;?)(q24;?), der(11)t(11;?)(?->::11p15->11q22.1::11q13->22.1:), der(12)t(12;?)(p11;?), 14p+, t(15;17)(q22;q11-12.1), der(19)t(10;19)(q21.1;p13.3)x2^a^ |
| U937 | 63(58-69)<3n>XXY, -2, -4, -6, +7, -9, -20, -21, +3mar, t(1;12)(q21;p13), der(5)t(1;5)(p22;q35), add(9)(p22), t(10;11)(p14;q23), i(11q), i(12p), add(16)(q22), add(19)(q13)^a^ |
| MV4-11 | 48,XY, t(4;11)(q21;q23), +8, +19^b^ |
| SKM1 | 43(38-43)<2n>XY, +1, -12, -14, -20, -21, t(1;19)(q21;q13), del(2)(p11), del(9)(q12), add(17)(p1?)^a^ |
| ^a^ Data from <https://www.dsmz.de/collection/catalogue/human-and-animal-cell-lines/catalogue>  ^b^ Data from <https://www.lgcstandards-atcc.org/products/all/CRL-9591.aspx?geo_country=gb#characteristics> | |

**Table S2. Significantly differentially methylated CpGs (*P* < 0.05) in HEL cell clones with biallelic *TET2* mutations compared to parental clones with monoallelic *TET2* mutations.**

(Excel file)

**Table S3. Significantly differentially expressed genes (*P*_adj_ < 0.05 and |Log_2_FC| ≥ 0.3) in HEL cell clones with biallelic *TET2* mutations compared to parental clones with monoallelic *TET2* mutation identified.**

(Excel file)

**Table S4. Gene ontology component analysis of significantly differentially expressed genes in HEL cell clones with biallelic *TET2* mutations compared to parental clones with monoallelic *TET2* mutation.**

(Excel file)

**Table S5. Significantly differential expression (*P*_adj_ < 0.05) of components of the snRNP complex (GO:0097525) in HEL cell clones with biallelic *TET2* mutations compared to parental clones with monoallelic *TET2* mutation.**

(Excel file)

**Table S6. Gene ontology biological pathway analysis of significantly differentially expressed genes in HEL cell clones with biallelic *TET2* mutations compared to parental clones with monoallelic *TET2* mutation.**

(Excel file)

**Table S7. Clinical characteristics and demographics of AML patients with a cytogenetically discernible chromosome 4 aberration (from SAL AML Biobank).**

| **Patient ID** | **Gender** | **Karyotype** | **Copy number alteration affecting 4q24 (from cytogenetics)** | **Age at diagnosis** | **ELN risk^a^** | **WBC count** | **Treatment^b^** | **CR** | **ALL-SCT^c^** | **Relapse** | **OS (months)** | **Status**  **(at last follow-up)** | ***TET2* copy number alteration (from SNP array)** | ***TET2* base substitution (from Sanger sequencing)** | ***TET2* loss of function mutation status** |
| --- | --- | --- | --- | --- | --- | --- | --- | --- | --- | --- | --- | --- | --- | --- | --- |
| UPN01  (index case) | M | 46,XY,t(4;12)(q2?;q13)[12]/46,XY[10] | not discernible | 76-80 | int | 38.2 | DA1, AZA | no | - | yes | 27.9 | deceased | focal monoallelic deletion | c.2815C>T, p.Gln939* | biallelic |
| UPN02 | M | 42~44,X,-Y,der(4)t(4;12)(q21;q24),del(5)(q13q33),-6,-11,der(12)t(6;12)(q21p12),t(12;6)(q13;q21),t(6;4)(q24;q25),-13,der(15)t(11;15)(q21;p11),t(11;15)(q21;q23),der(16)t(13;16)(q14;q12),-18,der(19)t(19;20)(q12;q12),der(20)t(15;20)(q23;q11)[cp19]/46,XY[1] | negative | 46-50 | adv | 6.9 | DA1 | yes | salvage | yes | 22.8 | deceased | negative | negative |  |
| UPN03 | F | 46,XX,der(4)del(4)(q?11)t(4;?11)(q?11;q23q23),der(7)del(7)(q?11)t(7;?10)(q?11;p12),+8,der(10;11)(10qter-->10p12::11q23-->11q23:?11p11-->11q13::4q?21-->4qter)[8] | negative | 21-25 | adv | 14.3 | DA1 | yes | salvage | yes | 26.5 | deceased | negative | negative |  |
| UPN04 | M | 46,XY,der(3)inv(3)(q21q26)add(3)(p25),t(4;6)(q35;q23),del(7)(q21q31),t(8;21)(q22;q22),add(14)(q32.1),add(15)(q24)[11]/45,X,-Y,der(3)inv(3)(q21q26)add(3)(p25),t(4;6)(q35;q23),del(7)(q21q31),t(8;21)(q22;q22),add(14)(q32.1),add(15)(q24)[9] | negative | 26-40 | fav | 21.9 | DA1, DA2 | yes | post-CR | yes | 76.8 | alive | negative | negative |  |
| UPN05 | F | 46,XX,t(4;22)(q25;q13)[5]/46,XX[20] | negative | 56-60 | int | 25.8 | DA1, DA2, MAC, MAMAC | yes | salvage | yes | 12.6 | deceased | negative | negative |  |
| UPN06 | M | 42,XY,+der(1)t(1;16),del(2),?der(3)t(3;15),?+del(4),-5,der(7)t(7;22),der(7)t(2;7;16),?i(8q),-9,der(12)t(5;12),-15,-16,-17,t(17;18),-18,-22,?ins(22;3),+mar[3]/42,XY,+der(1)t(1;16),del(2),der(3)t(3;15),?+del(4),-5,-6,der(7)t(2;7;16),der(12)t(5;12),-13,-13, | not discernible | 51-55 | adv | 4.1 | DA1 | yes | salvage | yes | 6.4 | deceased | negative | negative |  |
| UPN07 | M | 38-45,XY,del(1)(?q21),ins(1;4)(q?21;?),der(3)(3qter->3p21::?17q11.2->?17q21::?6->?6),t(5;17)(p10;q10),del(6)t(6;12)(p21;?),der(6)t(6;17)(p21;?),del(7)t(7;17)(p13;?)t(?3;7)(p21;q?22),+8,i(8)(q10)+1-2,-16,-17[cp14]/46,XY[1] | not discernible | 51-55 | adv | 43.24 | DA1 | no | - | no | 2.4 | deceased | negative | negative |  |
| UPN08 | M | 46,XY,der(5)del(5)(q31)t(4;5)(?;3)[2]/46,XY,der(5)del(5)(q31)t(4;5)(?;q31)[6] | not discernible | 16-20 | adv | 34.2 | DA1, DA2 | no | - | no | 1.6 | deceased | negative | negative |  |
| UPN09 | M | 46-49,XY,del(4)(q13),add(7)(q11),der(17;18)(q10;q10),der(21)t(14;21)(q12;q11),+mar1-3[cp16]/46-49,XY,+19,der(21)t(14;21)q22;q11),+22[cp4] | negative | 51-55 | adv | 3.7 | DA1 | yes | - | no | 1.2 | deceased | large monoallelic deletion (*TET2* copy number < 2) | negative | monoallelic |
| UPN10 | F | 46,XX,add(4)(q31),del(4)(q24q28),-5,del(7)(q21q35),add(12)(p11),-17,-20,+1-2mar[13]/45,idem,-3,del(4)(q24q28),del(8)(q12q13),-add(12)(p11,add(18)(q12) [2] | deletion 4q24 | 46-50 | adv | 4.6 | DA1 | yes | post-CR | no | 100.6 | alive | large monoallelic deletion (*TET2* copy number < 2) | negative | monoallelic |
| UPN11 | M | 47,XY,del(3)(p14),der(4)t(4;5)(q26;?),der(5)t(5;12)(q14;?),der(6;21)(q10;q10),der(6)ins(6;21)(q11;q11q22),t(6;21)(p25;q11),der(7)t(7;12)(q11;q?),der(12)t(7;12)(?;q21),t(4;7)(?;?),r(21)ins(21;18),+r(21)ins(21;18)[cp13]/46,XY[3] | not discernible | 51-55 | adv | 2.41 | DA1 | yes | salvage | yes | 4.7 | deceased | negative | negative |  |
| UPN12 | M | 47,XY,+21[12]/47,XY,+21,del(4)(q?)[3]/47,XY,del(13)(q?)[5]/47,XY,+21,del(4)(q?),del(13)(q?)[1] | not discernible | 41-45 | fav | 277 | DA1, DA2 | yes | salvage | yes | 131.8 | alive | negative | negative |  |
| UPN13 | F | 47~49,XX,del(1)(p13p22),-4,del(5)(q13q33),+8,+8,add(11)(q2?3),der(12)t(4;12)(q21;p1?3),-13,-14,?der(16),-17,-18,-19,+3-7mar[cp10] | monosomy 4 | 71-75 | adv | 9 | DA1 | no | - | no | 0.3 | deceased | negative | negative |  |
| UPN14 | M | 46,XY,t(4;14)(q11;q32)[12]/46,XY[1] | negative | 61-65 | int | 5.9 | DA1, DA2, MAMAC | yes | - | yes | 66.7 | deceased | negative | negative |  |
| UPN15 | M | 46,XY,t(4;21)(q11;q11)[23] | negative | 31-35 | int | 21.7 | MAV, MAMAC | yes | post-CR | no | 94.4 | alive | negative | negative |  |
| UPN16 | F | 45,XX,?t(4;10)(p11;p15),del(5)(q13q33),-7,del(12)(p13)[15] | negative | 61-65 | adv | 43 | DA1 | no | - | no | 4.6 | deceased | negative | negative |  |
| UPN17 | M | 45~52,XY,der(1)t(1;3)(p22;p11),der(3)t(1;3;5)(p22;p11;q11),del(6)(q21),?del(8)(q22),-12,del(13)(q13q22),der(20)t(4;20)(q13;q11),+mar,inc[cp30] | negative | 71-75 | adv | 2.1 | DA1 | no | - | no | 2.4 | deceased | negative | negative |  |
| UPN18 | M | 43~44,XY,del(1)(q32),der(3)(p?),del(4)(q?),del(5)(q13q31),-7,add(8)(p23),i(11)(q10),add(17)(q25),add(22)(q13),add(22)(q13),inc[cp27] | not discernible | 71-75 | adv | 28.9 | DA1 | no | - | no | 0.6 | deceased | large monoallelic deletion (*TET2* copy number < 2) | negative | monoallelic |
| UPN19 | F | 46,XX,der(4)(q31),del(9)(q13q22),-12,del(13)(q14q22),der(14)(q13),del(20)(q11),+mar[cp19]/46,XX [2] | negative | 61-65 | adv | 1.6 | DA1 | yes | - | yes | 21.3 | deceased | negative | negative |  |
| UPN20 | F | 46,XX,t(4;12)(q1?1;p1?2)[13]/46,XX[4] | negative | 61-65 | int | 3.08 | DA1, DA2 | no | - | no | 2.7 | deceased | negative | negative |  |
| UPN21 | F | 47,XX,+4[22] | trisomy 4 | 71-75 | fav | 9 | DA1, DA2, MAMAC | yes | - | yes | 6 | deceased | trisomy 4 plus LOH affecting most of the long arm, including *TET2* (copy number > 2) | negative |  |
| UPN22 | F | 47,XX,+4[8]/48,XX,+4,+4[7]/48,XX,+4,+8[4]/50,XX,+4,+8,+13,+19[1] | trisomy 4 | 66-70 | fav | 30.8 | DA1 | no | - | no | 0.8 | deceased | trisomy 4 | negative |  |
| UPN23 | M | 53,XY,+4,+6,+8,+9,add(17)(q25),+18,+21,+21[26]/46,XY[2] | trisomy 4 | 16-20 | adv | 5.3 | MAV | yes | post-CR | yes | 7.8 | deceased | trisomy 4 | negative |  |
| UPN24 | F | 49,XX,+4,+8,t(10;11)(p13;q14),+12[10] | trisomy 4 | 51-55 | adv | 171 | MAV | no | - | no | 0.8 | deceased | trisomy 4 | negative |  |
| UPN25 | M | 48,XY,+4,-6,+8,+mar[9]/46,XY[21] | trisomy 4 | 61-65 | adv | 14.5 | MAV, MAMAC | yes | - | yes | 32.9 | deceased | trisomy 4 plus LOH affecting most of the long arm, including *TET2* (copy number > 2) | c.4133G>A, p.Cys1378Tyr | biallelic |
| UPN26 | F | 47,XX,+4,t(8;21)(q22;q22)[9]/46,XX,t(8;21)(q22;q22)[18] | trisomy 4 | 51-55 | fav | 34.8 | MAV, MAMAC | yes | - | yes | 17.7 | deceased | negative | negative |  |
| UPN27 | F | 47,XX,+4[18]/46,XX[2] | trisomy 4 | 61-65 | fav | 1.2 | DA1, DA2 | yes | salvage | yes | 22.1 | deceased | trisomy 4 | negative |  |
| UPN28 | M | 47,XY,+4[30] | trisomy 4 | 71-75 | int | 36.3 | DA1 | yes | - | no | 62 | deceased | trisomy 4 with a focal deletion affecting *TET2* (copy number < 2) | negative | monoallelic |
| UPN29 | F | 46,XX,t(9;11)(p21-22;q23)[14]/49,XX,+4,+8,t(9;11)(p21-22;q23),+12[2] | trisomy 4 | 36-40 | adv | 212 | DA1 | no | - | no | 0.1 | deceased | negative | negative |  |
| UPN30 | F | 47,XX,+8[1]/47,der(3)del(3)(p13)ins(3;12)(q21;q13q24.1),del(4)(q2?),+8,der(12)del(12)(q13q24.1)ins(12;4)(q13;?)[21]/47,XX,idem,?add(13)(q34)[2]/46,XX[7] | not discernible | 71-75 | adv | 4.7 | none | no | - | no | 23.8 | deceased | focal monoallelic deletion | c.3646C>T, p.Arg1216* | biallelic |
| ^a^ Risk category according to European LeukaemiaNet (ELN). fav, favourable; int, intermediate; adv, adverse.  ^b^ Remission-induction treatment regime(s) administered. DA, daunorubicin and Ara-C; MAC, mitoxantrone and Ara-C; MAMAC, amsacrine and Ara-C; MAV, mitoxantrone, Ara-C and VP-16; AZA, 5’-Azacytidine  ^c^ Patients who received allograft stem cell transplant (ALL-SCT) either after achieving CR (post-CR) or as salvage therapy after relapse (salvage). | | | | | | | | | | | | | | | |

**Table S8. Clinical characteristics and demographics of AML patients with *TET2* mutation enrolled in the PETHEME FLUGAZA phase 3 clinical trial.**

| **Patient ID** | **Gender** | **Age at diagnosis** | **ECOG score^a^** | **ELN risk^b^** | **Treatment^c^** | **CR** | **OS (days)** | **Status (at last follow-up)** | **Cause of death** | ***TET2* loss of function mutation status** |
| --- | --- | --- | --- | --- | --- | --- | --- | --- | --- | --- |
| UPN01 (index case) | M | 76-80 | 2 | fav | DA, AZA | Yes | 850 | deceased | Progression | biallelic |
| UPN31 | M | 81-85 | 3 | adv | AZA | Yes | 767 | deceased | Progression | biallelic |
| UPN32 | M | 66-70 | 2 | fav | AZA | Yes | 588 | deceased | Progression | monoallelic |
| UPN33 | M | 66-70 | 3 | adv | AZA | Yes | 579 | deceased | Progression | biallelic |
| UPN34 | M | 71-75 | 1 | adv | AZA | No | 523 | deceased | Progression | monoallelic |
| UPN35 | M | 76-80 | 0 | adv | AZA | Yes | 517 | deceased | Progression | monoallelic |
| UPN36 | M | 81-85 | 2 | adv | AZA | No | 507 | deceased | Progression | monoallelic |
| UPN37 | F | 71-75 | 0 | int | AZA | No | 470 | deceased | Progression | monoallelic |
| UPN38 | M | 71-75 | 0 | int | AZA | Yes | 435 | deceased | Cardiovascular disease | monoallelic |
| UPN39 | F | 81-85 | 3 | adv | AZA | Yes | 373 | deceased | Progression | monoallelic |
| UPN40 | M | 66-70 | 1 | adv | AZA | Yes | 356 | deceased | Progression | monoallelic |
| UPN41 | M | 76-80 | 1 | fav | AZA | Yes | 344 | deceased | Progression | monoallelic |
| UPN42 | M | 66-70 | 2 | fav | AZA | No | 183 | deceased | Progression | monoallelic |
| UPN43 | F | 86-90 | 2 | int | AZA | No | 125 | deceased | Infection | monoallelic |
| UPN44 | M | 76-80 | 0 | adv | AZA | No | 114 | alive | - | monoallelic |
| UPN45 | M | 71-75 | 1 | adv | AZA | No | 106 | deceased | Progression | monoallelic |
| UPN46 | F | 76-80 | 1 | adv | AZA | No | 82 | deceased | Cardiovascular disease | monoallelic |
| UPN47 | M | 76-80 | 3 | adv | AZA | No | 62 | deceased | Progression | biallelic |
| UPN48 | M | 71-75 | 1 | adv | AZA | NA | 56 | deceased | Infection | monoallelic |
| UPN49 | M | 71-75 | 2 | adv | AZA | No | 52 | deceased | Hemorrhagic complication | monoallelic |
| UPN50 | F | 86-90 | 3 | adv | AZA | No | 26 | deceased | Progression | monoallelic |
| UPN51 | M | 66-70 | 1 | int | FLUGA | Yes | 1308 | alive | - | monoallelic |
| UPN52 | M | 71-75 | 0 | int | FLUGA | No | 1023 | deceased | Progression | monoallelic |
| UPN53 | M | 66-70 | 0 | int | FLUGA | No | 862 | deceased | Progression | monoallelic |
| UPN54 | M | 76-80 | 1 | adv | FLUGA | No | 620 | deceased | Lung cancer | monoallelic |
| UPN55 | F | 71-75 | 2 | int | FLUGA | Yes | 551 | deceased | Progression | monoallelic |
| UPN56 | M | 76-80 | 1 | fav | FLUGA | Yes | 473 | deceased | Hemorrhagic complication | monoallelic |
| UPN57 | M | 66-70 | 1 | fav | FLUGA | Yes | 420 | deceased | Progression | monoallelic |
| UPN58 | F | 76-80 | 0 | adv | FLUGA | Yes | 388 | deceased | Progression | monoallelic |
| UPN59 | M | 76-80 | 0 | adv | FLUGA | Yes | 362 | deceased | Possible stroke | monoallelic |
| UPN60 | F | 76-80 | 1 | adv | FLUGA | Yes | 354 | deceased | Progression | monoallelic |
| UPN61 | F | 66-70 | 1 | int | FLUGA | Yes | 267 | deceased | Progression | monoallelic |
| UPN62 | F | 71-75 | 1 | adv | FLUGA | Yes | 220 | deceased | Progression | monoallelic |
| UPN63 | M | 71-75 | 0 | fav | FLUGA | No | 203 | deceased | Progression | monoallelic |
| UPN64 | F | 61-65 | 1 | int | FLUGA | No | 159 | deceased | Progression | monoallelic |
| UPN65 | M | 76-80 | 1 | adv | FLUGA | No | 155 | deceased | Progression | monoallelic |
| UPN66 | F | 66-70 | 1 | int | FLUGA | No | 141 | deceased | Progression | monoallelic |
| UPN67 | F | 81-85 | 1 | adv | FLUGA | No | 126 | deceased | Progression | monoallelic |
| UPN68 | M | 71-75 | 0 | adv | FLUGA | No | 111 | deceased | Progression | biallelic |
| UPN69 | F | 66-70 | 2 | adv | FLUGA | No | 100 | deceased | Progression | monoallelic |
| UPN70 | M | 66-70 | 2 | adv | FLUGA | No | 94 | deceased | Infection | monoallelic |
| UPN71 | M | 76-80 | 0 | int | FLUGA | No | 68 | deceased | Progression | monoallelic |
| UPN72 | M | 81-85 | 0 | int | FLUGA | NA | 53 | deceased | Myocardial infarction | monoallelic |
| UPN73 | F | 71-75 | 2 | int | FLUGA | No | 45 | deceased | Progression | biallelic |
| UPN74 | F | 76-80 | 1 | adv | FLUGA | No | 40 | deceased | Infection | monoallelic |
| UPN75 | F | 81-85 | 1 | adv | FLUGA | Yes | 37 | deceased | Infection | monoallelic |
| UPN76 | M | 81-85 | 1 | adv | FLUGA | NA | 31 | deceased | Infection | monoallelic |
| UPN77 | F | 71-75 | 0 | fav | FLUGA | NA | 19 | deceased | Hemorrhagic complication | monoallelic |
| UPN78 | M | 71-75 | 1 | int | FLUGA | NA | 17 | deceased | Hemorrhagic complication | biallelic |
| UPN79 | F | 76-80 | 1 | fav | FLUGA | NA | 9 | deceased | Progression | monoallelic |
| UPN80 | M | 81-85 | 1 | adv | FLUGA | No | 3 | deceased | Tumor lysis syndrome | monoallelic |
| ^a^ Performance status according to European Cooperative Oncology Group (ECOG).  ^b^ Risk category according to European LeukaemiaNet (ELN). fav, favourable; int, intermediate; adv, adverse.  ^c^ Remission-induction treatment regime(s) administered. DA, daunorubicin and Ara-C; AZA, 5’-Azacytidine; FLUGA, low-dose Ara-C and fludarabine | | | | | | | | | | |

**Table S9. Primer sequences and PCR reaction conditions for Sanger sequencing of genes of interest.**

| **Amplicon** | **Primer sequences^a^** | **PCR reaction conditions^b^** |
| --- | --- | --- |
| *TET2* exon 3 | F 5'-GCTTTCAAGAACAGGAGCAGA-3' *  R 5'-CAGGCATGTGGCTTGCATC-3' | 35 cycles:  D 95°C 25s  A 60°C 35s  E 72°C 45s |
| *TET2* exon 6 | F 5’-TGCAAGTGACCCTTGTTTTG-3’ *  R 5’-TACCGAGACGCTGAGGAAAT-3’ | 36 cycles:  D 95°C 25s  A 55°C 35s  E 72°C 65s |
| *NPM1* exon 11 | F 5’-AACTCTCTGGTGGTAGAATGAAA-3’  R 5’-TGAGAACTTTCCCTACCGTGT-3’ * | 36 cycles:  D 95°C 25s  A 55°C 35s  E 72°C 65s |
| *FLT3* exon 14 | F 5’-ACAGGGACATTGCCTGATTGT-3’  R 5’-GGTTGACACCCCAATCCACT-3’ * | 35 cycles:  D 95°C 25s  A 60°C 35s  E 72°C 45s |
| ^a^ F, forward; R, reverse.  ^b^ initial denaturation was performed at 95°C for 2 min and the last cycle was followed by a final extension step at 72°C for 5 min in all cases. D, denaturation; A, annealing; E, extension.  * indicates primer used for Sanger sequencing. | | |
